# Human Organ Chips Reveal New Inflammatory Bowel Disease Drivers

**DOI:** 10.1101/2024.12.05.24318563

**Authors:** Alican Özkan, Gwenn E. Merry, David B. Chou, Ryan R. Posey, Anna Stejskalova, Karina Calderon, Megan Sperry, Joshua Piatok, Viktor Horvath, Lorenzo E. Ferri, Emanuela Carlotti, Stuart A. C. McDonald, Douglas J. Winton, Rocco Riccardi, Liliana Bordeianou, Sean Hall, Girija Goyal, Donald E. Ingber

**Author notes:** Address correspondence to: Donald E. Ingber, MD, PhD 201 Brookline Ave., Suite 401, Boston MA, 02215, USA em.

## Abstract

Inflammatory bowel disease (IBD) patients exhibit compromised intestinal barrier function and decreased mucus accumulation, as well as increased inflammation, fibrosis, and cancer risk, with symptoms often being exacerbated in women during pregnancy. Here, we show that these IBD hallmarks can be replicated using human Organ Chips lined by IBD patient-derived colon epithelial cells interfaced with matched fibroblasts cultured under flow. Use of heterotypic tissue recombinants revealed that IBD fibroblasts are the primary drivers of multiple IBD symptoms. Inflammation and fibrosis are accentuated by peristalsis-like motions in IBD Chips and when exposed to pregnancy-associated hormones in female IBD Chips. Carcinogen exposure also increases inflammation, gene mutations, and chromosome duplication in IBD Chips, but not in Healthy Chips. These data enabled by human Organ Chip technology suggest that the intestinal stroma, sex hormones, and peristalsis-associated mechanical deformations play a key role in driving inflammation, fibrosis, and disease progression in male and female IBD patients.

## Main

Inflammation of the intestinal mucosa, including enhanced production of inflammatory cytokines and tissue infiltration with immune cells, along with increased barrier permeability, thinning of the protective mucus layer, and fibrosis are central hallmarks of inflammatory bowel diseases (IBD), such as Crohn’s disease (CD) and ulcerative colitis (UC). These alterations lead to significant digestive issues and an increased risk of developing colorectal cancer^1^. IBD symptoms also can be more prominent in women^2^ and they have to control their symptoms before pregnancy because many of these patients experience exacerbations that increase the risk of preterm birth^3^. Most IBD treatments target immune cells rather than intestinal tissues, and treatment efficacy is variable among patients^4^. However, interactions between epithelia and underlying stroma also may contribute to inflammation as well as cancer progression^5^. For example, the presence of a subset of fibroblasts that recruit immune cells to the intestinal stem cell niche is associated with poor prognosis in patients with colorectal cancer^6^. Inflamed epithelial cells also can undergo an epithelial-to-mesenchymal transition and increase deposition of extracellular matrix (ECM) resulting in intestinal fibrosis that leads to complications, such as bowel stiffening and restriction of lumenal fluid flow, which further exacerbate digestive problems in IBD patients ^7,8^. Thus, there is a need to gain greater insight into how epithelial-stromal interactions, peristalsis, and fluid flow influence IBD development in both male and female patients. However, animal IBD models and in vitro studies with human intestinal cell lines lack direct relevance to the human disease^9^, and human intestinal organoids cannot be used to address these questions because they lack an epithelial-stromal interface, mucosal barrier, fluid flow, peristalsis motions, and immune cells, which are critical organ-level features required to address these questions.

Here, we leveraged human organ-on-a-chip (Organ Chip) microfluidic culture technology that recreates tissue-tissue interfaces and a physiologically relevant intestinal microenvironment including dynamic fluid flow, peristalsis-like mechanical deformations, and circulating immune cells to confront this challenge^10,11^. Colon Chips lined by human organoid-derived colonic epithelium cultured under dynamic fluid flow have been successfully used in the past to study mucus layer accumulation and physiology^12,13^ as well as to identify microbiome metabolites that influence host response to bacterial infections^14^. In this study, we isolated epithelium and stromal-derived fibroblasts from the same regions of colon from healthy or IBD patients and used them to create Colon Chips that contained an epithelial-fibroblast interface. We also created heterotypic tissue recombinant chips, measured the effects of applying peristalsis-like motions and flowing immune cells through the stromal channel, and studied the effects of female hormones on the IBD state, in addition to modeling cancer progression by exposing the Colon Chips to carcinogens in vitro.

## Results

### Establishment of Human Healthy and IBD Colon Chips

Patient-specific epithelial organoids and stromal fibroblasts were isolated from surgical resections of the colon of healthy and IBD patients with CD or UC (**Extended Data Fig. 1A** and **Supplementary Information**) and the organoids were expanded in Matrigel cultures while the fibroblasts were cultured on plastic dishes (**Extended Data Fig. 1B**). To build the human Colon Chips, the primary colon epithelial cells and fibroblasts were respectively seeded on the top and bottom surfaces of a porous ECM-coated membrane that separates two parallel channels within a commercially available microfluidic Organ Chip (**Fig. 1A**). The cells were allowed to adhere under static conditions for 1 day before flow was initiated. A continuous epithelial monolayer with undulating crypt-like structures formed by 11 days in Healthy Chips when viewed from above (**Fig. 1B,C)** or in cross sections (**Fig. 1D,E**). The epithelium became progressively obscured by dense opaque deposits from day 3 to 7, which completely filled the upper channel by day 11 (**Fig. 1C**). These opaque materials have been shown to be composed of mucus with similar composition, thickness, and bilayer structure as seen in human colonic mucus in vivo ^12,13^. Similar chips lined with colonic epithelium and fibroblasts from CD or UC patient samples also formed hill and crypt-like structures, but significantly less mucus accumulated on top of the epithelium, particularly in the UC chips (**Fig. 1C** and **Extended Data Fig. 1C**). Analysis of vertical cross-sectional views of the intestinal epithelium cultured in chips revealed that the heights of the undulating epithelial layerwere significantly reduced in the IBD chips as well (**Fig. 1D and Extended Data Fig. 1D**). Epithelial cells within both Healthy and IBD Chips also expressed colon-specific cytokeratin 20 along their apical borders, while the basal stem cell marker SOX9 primarily appeared in their nuclei at the cell base (**Fig. 1F**), which is a hallmark of colon crypts in vivo ^15^.

**Figure 1:**
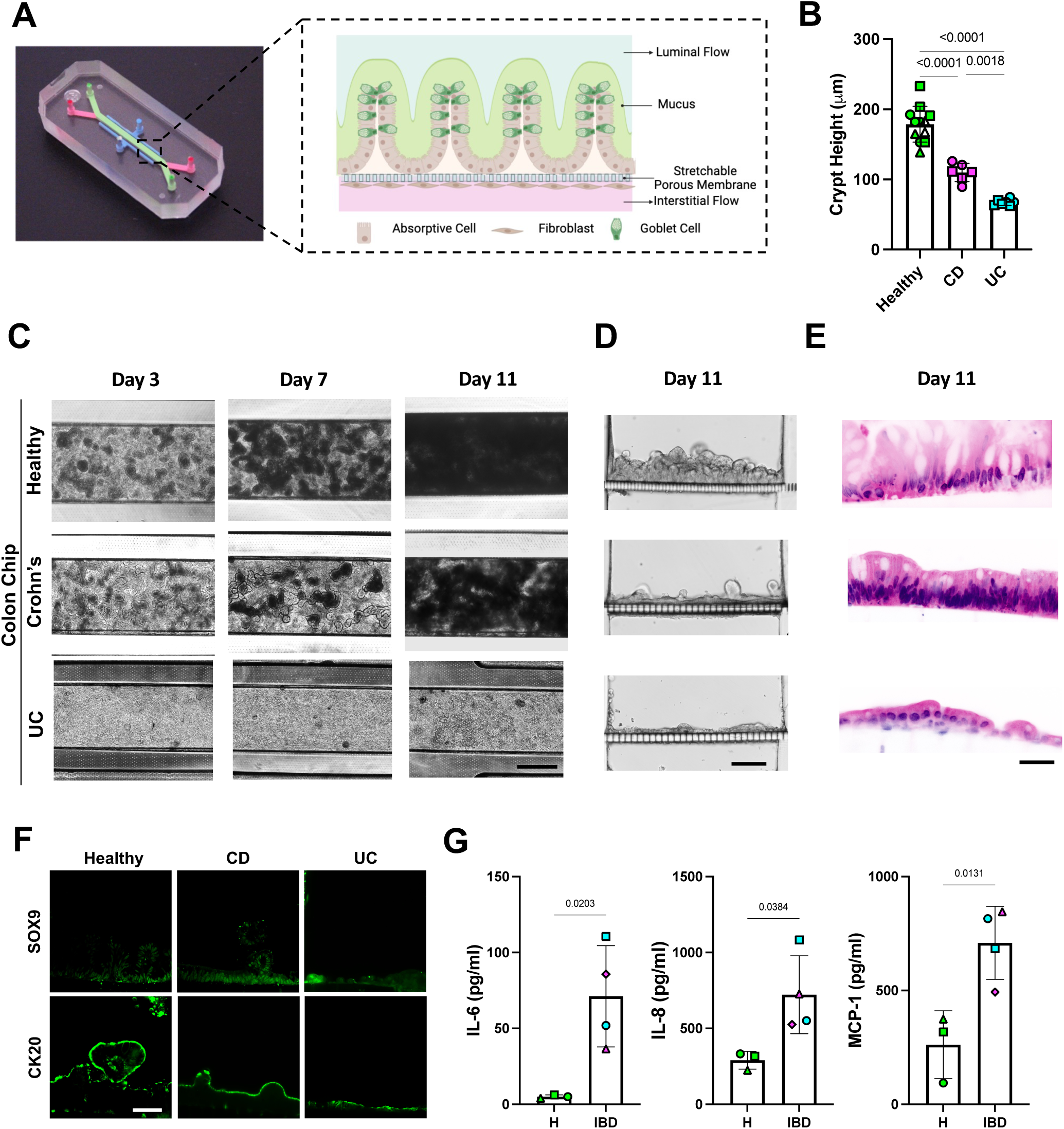
Primary human Colon Chips generated with epithelium and fibroblasts isolated from Healthy and IBD patient samples. **A)** Photograph of the commercially available Organ Chip device (left) and a cross-sectional illustration of the epithelial-stromal tissue interface (right) formed with patient-derived cells, which is composed of a fibroblast rich stroma that is perfused with medium to mimic interstitial fluid flow separated by a flexible, porous, ECM-coated membrane from a colon epithelium above. The epithelium, which is composed of absorptive cells and Goblet cells, secretes a thick mucus layer into the lumen of the apical channel that also experiences fluid flow. The entire tissue-tissue interface can be stretched and relaxed rhythmically to mimic peristalsis-like motions by applying cyclic suction to hollow side chambers in the flexible device (not shown). **B)** Height of crypts formed in Colon Chips measured at 11 days. Numbers indicate *P* values between compared groups, as determined by one-way ANOVA test (n=3 Healthy, green; 2 CD, magenta; and 2 UC, cyan). **C)** Representative bright field microscopic images of Healthy and IBD Chips created with cells from CD or UC patients viewed from above on day 3, 7, and 11. A functional epithelial monolayer progressively accumulated mucus, which appears as opaque blackened fuzzy material, in the apical channel of all chips over the 11 day time course; however, mucus accumulation was suppressed in the IBD Chips with the UC Chips displaying a greater reduction (bar, 500 µm)**. D)** Brightfield image of vertical cross-sections through healthy and IBD Colon chips showing that the crypt-like epithelial structures are shorter in the CD and UC Colon Chips (bar, 100 µm)**. E)** Histological H&E stained vertical cross-sections through healthy and IBD Colon chips confirming the presence of columnar colon epithelium containing goblet cells in healthy epithelium, and a significant reduction in the height of the epithelial monolayer with reduced numbers of goblet cells in the IBD Chips (bar, 50 µm). **F)** Immunofluorescence microscopic views showing distribution of SOX9 and CK20 in crypts in the colon epithelium of Healthy and IBD Chips (bar, 50 µm). **G**) Inflammatory cytokine protein levels measured in the basal outflows of Healthy (H) and IBD Chips on culture day 11 (n = 3-5 chips/ condition; chips were created with cells from 3 healthy (green), 2 CD (magenta), and 2 UC (cyan) patient donors, with each symbol represents a chip created with cells from a different patient. Numbers indicate *P* values between compared groups, as determined by two-tailed student’s t-test. All data represent mean ± SD; *p* values are shown in all figures. Source data and statistical tests are provided as a Source Data file.

Past work has shown that there is a subset of stromal fibroblasts in IBD patients that have higher levels of expression of oncostatin M receptor (OSMR) and podoplanin (PDPN)^6^, which appear to play a significant role in inflammation, fibrosis, and cancer^16^. Indeed, when we carried out flow cytometric analysis after removing stromal cells from the chips, we found that PDPN expression was significantly higher in IBD fibroblasts that expressed high OSMR levels (**Extended Data Fig. 2A,B**) and that fibroblasts in IBD Chips lined by cells from both CD and UC patients secreted more inflammatory cytokines (e.g, IL6, IL8, MCP-1) from the basal channel than those in Healthy Chips (**Fig. 1G**). We also found that fibroblasts in IBD Chips established using both CD and UC patients samples express higher vimentin and α-SMA than healthy controls (**Extended Data Fig. 2C,D**), as previously observed in IBD mice models induced by treatment with DSS or IL10 knock out^17^.

Transcriptomic analysis of epithelial cells and fibroblasts individually within Healthy versus IBD Chips revealed that genes which were found to be overexpressed in CD and UC (e.g., ADA2, APOBEC3B, GSPT2, CD40, ACAN, HLA-DQB1) epithelium on-chip were also expressed at higher levels in the intestinal epithelium of IBD patients^18–22^ compared to healthy patients (**Extended Data Fig. 3A,B**). In addition, genes suppressed in IBD patient tissues in vivo (e.g., MUC5B, FDFT1 in CD and SOD3, SMIM32, APOBR in UC ^18–22)^ were expressed at lower levels by epithelium in both CD and UC IBD Chips.

When stromal cells cultured in IBD Chips were similarly analyzed, they displayed expression of IBD-relevant genes with CD fibroblasts overexpressing ITGA7, FXD1, HGF, FZD7, NKX2-3, LOXL3 relative to healthy fibroblasts, while high levels of IKBIP, NKX2-3, ADA2, ECM2, NDN, SLC22A17 were seen in UC fibroblasts **(Extended Data. Fig. 3C,D**). In contrast, other genes were downregulated in these fibroblasts including NLRP2, FUCA1, CLCA4, and RNF186 in CD and RALGPS1, DLG3, STON2, and FMO5 in UC derived IBD Chips (**Extended Data Fig. 3C,D**), which is again consistent with results of transcriptomic analyses carried out on IBD patient tissues^23–25^.

Gene ontology and Reactome analysis revealed that multiple IBD-relevant biological processes, including increased ECM production and remodeling, cell adhesion, and angiogenesis as well as decreased metabolism were predominant in both CD and UC epithelium and fibroblasts on-chip, with UC tissues also exhibiting suppressed IL10 signaling (**Extended Data Fig. 4A-D**). These functional pathways are also activated or suppressed in a similar manner in IBD patients^26^.

### Peristalsis-Like Mechanical Motions Enhance IBD Progression

IBD patients experience thinning of the protective mucus layer in their colon^12^ and eventually loss of peristalsis as a consequence of increased fibrosis^7,8^, which further exacerbate their gastrointestinal symptoms. However, it is not known whether cyclic mechanical deformation of inflamed intestine or fluid flow due to peristalsis could influence IBD disease progression. To explore this possibility, we cultured Healthy and IBD Chips in the presence or absence of physiologically relevant cyclic deformations by applying suction to side chambers in the flexible devices. The healthy and IBD epithelia both accumulated a mucus layer when exposed to cyclic mechanical deformations were viewed by dark field imaging; however, the layer was much thinner in the IBD Chips (**Fig. 2A,B**). Transcriptomic analysis confirmed that the IBD Chips expressed lower levels of most mucins than Healthy Chips under all conditions, and that while expression of MUC-1,4,12,13,17,3A,5AC, and 5B increased in Healthy Chips exposed to mechanical strain, only MUC5AC and MUC13 increased in the IBD Chips (**Fig. 2C)**. These data show that peristalsis-like mechanical deformations have a direct effect on mucus production in the colonic epithelium in both healthy and IBD Chips as fluid flow was constant under all conditions, and that the IBD epithelium is less effective at maintaining the normal thickness of the mucus layer in vitro as observed in IBD patients in vivo^12,13^.

**Figure 2:**
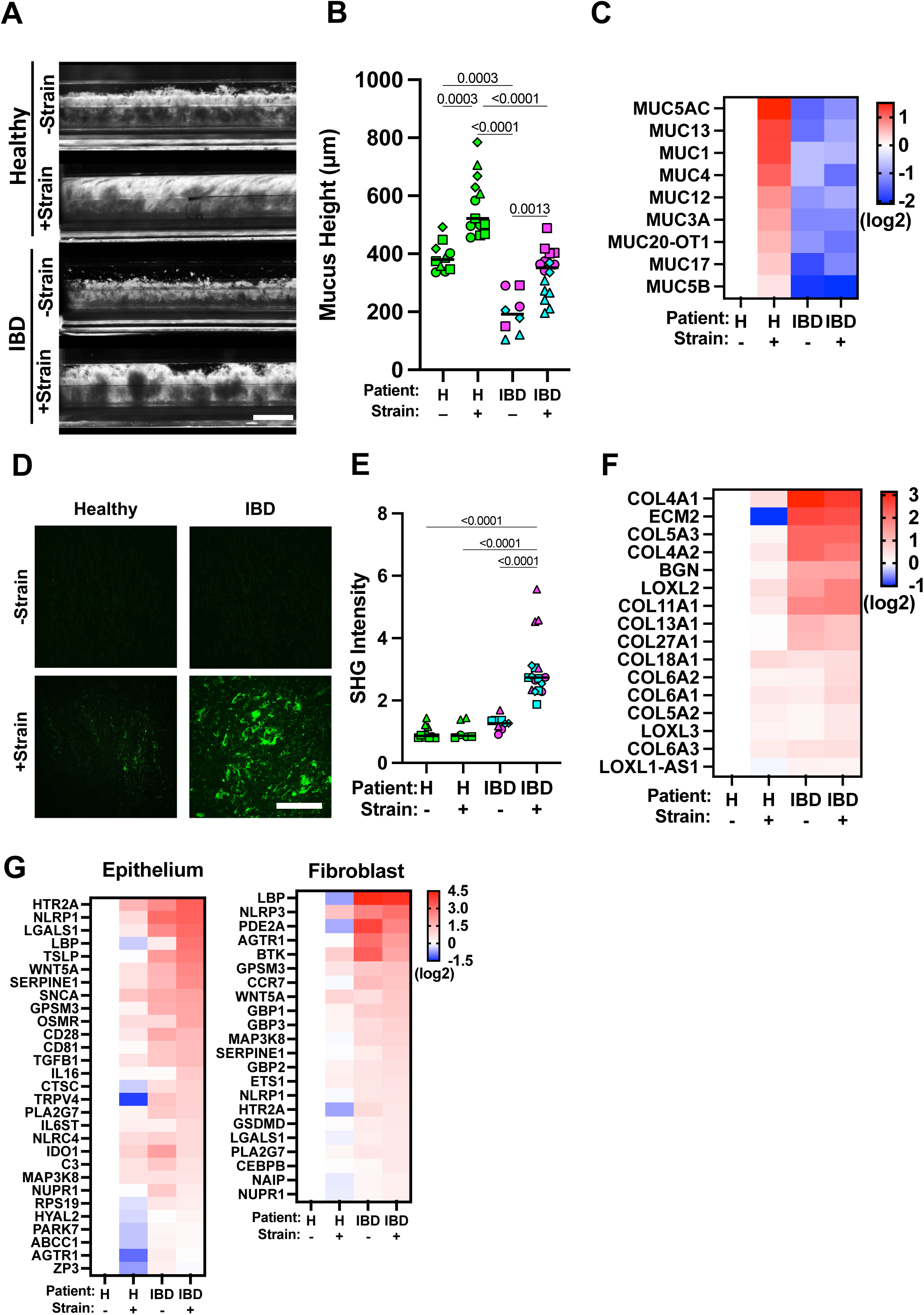
Peristalsis-like cyclic strain influences mucus production and fibrosis in Colon Chips. **A)** Representative side-view images microscopy of Healthy or IBD Chips with (+) and without (-) exposure to mechanical deformations visualizing mucus layer accumulation (white diffuse material) in Healthy Chips using dark-field microscopy. Note that limited mucus was produced by IBD epithelium (bar, 1 mm). **B)** Quantification of the height of the mucus layer overlying Healthy and IBD epithelium on-chip when cultured under the conditions in A. Numbers indicate *P* values between compared groups, as determined by one-way ANOVA test. (n = 3 healthy, green; 2 CD, magenta; and 2 UC, cyan). **C)** Heat map of genes showing that peristalsis-like mechanical stimulation (+) increases expression of genes related to mucus production in Healthy Chips, but not IBD Chips (n = 3 healthy, 1 CD and 2 UC). **D)** Second harmonic microscopic images of the fibroblast stroma in Healthy versus IBD Chips showing greater collagen fibril accumulation (green) with mechanical strain (+ Strain) in both chips, and that this response is greatly accentuated in IBD Chips (bar, 200 μm). **E)** Collagen fibril signal intensity in the stroma of Healthy versus IBD Chips quantified in second harmonic microscopic images. Numbers indicate *P* values between compared groups, as determined by one-way ANOVA test (n = 3 healthy, green; 2 CD, magenta; and 2 UC, cyan). **F)** Heat map of genes showing that peristalsis-like cyclic strain enhances expression of genes associated with fibrillar collagen production in IBD fibroblasts, but not in healthy fibroblasts (n = 3 healthy, 1 CD and 2 UC). **G)** Heat maps of showing that cyclic strain enhances greater expression of IBD-associated inflammatory genes in IBD Chips compared to Healthy Chips both in epithelium and fibroblasts (n=3 Healthy, 1 CD, and 2 UC). Source data and statistical tests are provided as a Source Data file.

Interestingly, second harmonic imaging of ECM fiber formation on-chip revealed that while both CD and UC derived IBD fibroblasts accumulate comparable fibrillar collagen as healthy cells, these levels increase many fold when the IBD Chips are exposed to peristalsis-like motions whereas there is no change in Healthy Chips (**Fig. 2D,E**). Atomic force microscopic analysis carried out in parallel samples from the resections we used to isolate the patient-derived cells confirmed that this was associated with tissue stiffening (**Extended Data Fig. 5A-D**). But while transcriptomic analysis showed that there is increased expression of collagen-associated genes in CD and UC stroma on-chip without mechanical strain, exposure to peristalsis-like deformations did not result in significant changes in collagen gene expression (**Fig. 2F**), suggesting that the fibrotic changes we observed resulted from post-transcriptional alterations. Indeed, pathway analysis confirmed that biological processes associated with fibrosis per se (ECM organization, collagen formation and fibril organization) were upregulated in IBD Chips upon exposure to cyclic mechanical deformations (**Extended Fig. 6A**). In addition, our transcriptomic analysis revealed that expression of different cytochrome P450 subunits was suppressed in IBD versus Healthy Chips, which is consistent with the observation that the intestinal tissues of IBD patients experience decreased metabolic activities^27,28^. Importantly, while peristalsis-like cyclic deformations increased the expression of most of these metabolic genes in Healthy Chips, this response was not observed in IBD Chips (**Extended Data Fig. 6B**).

Peristalsis-like cyclic deformations also increased barrier permeability in both Healthy and IBD Chips; however, barrier leakiness always remained higher in IBD Chips (**Extended Data Fig. 7A**). Mechanical stimulation also enriched expression of inflammation associated genes in epithelial cells and fibroblasts in both Healthy and IBD Chips (**Fig. 2G**) and this was supported by transcriptomic analysis, which showed activation of the inflammatory response pathway in these cells (**Extended Data Fig. 7B**). Importantly, further pathway analysis of this transcriptomic dataset (**Extended Data Fig. 8**) demonstrated that peristalsis-like deformations activate pathways associated with IBD progression and exacerbation in the epithelium, including those involved in cancer, Toll-like receptor signaling, PPAR signaling, and focal adhesion in IBD epithelium (**Extended Data Fig. 9A**) as well as NOD-like receptor signaling, PDGF signaling, and cytokine-cytokine receptor interaction in IBD fibroblasts (**Extended Data Fig. 9B**). In contrast, these pathways that are known to exacerbate colitis in IBD patients^29,30^ were not activated in Healthy Chips under the same mechanical stimulation conditions.

### Replicating Sex-Specific Clinical Symptoms of IBD Patients

The gastrointestinal symptoms of female IBD patients become much more prominent during menstrual periods^2^ and their therapies need to be optimized to control their symptoms before pregnancy due to increased risk of miscarriage, preterm birth and low birthweight^3^. Yet, it is not known whether these responses are a direct effect of ovarian hormones on the intestine, which could potentially contribute to the clinical observations observed in patients ^31^. To address this question directly, we exposed healthy and IBD Chips lined by cells from female patients to estrogen (E2) and medroxyprogesterone acetate (MPA) to recapitulate the menstrual cycle or a mixture of pregnancy-associated hormones (E2, MPA, human chorionic gonadotropin, prolactin, and placental lactogen) to mimic the first trimester of pregnancy. Interestingly, exposure to E2 and MPA or the pregnancy-associated hormone mixture, increased the height of crypt-like epithelial structures both in healthy and IBD chips (**Fig. 3A-C**). We also discovered addition of E2 and MPA increased fibrosis and exposure to the pregnancy-associated hormone mixture produced an even greater increase in fibrillar collagen deposition in the stroma of IBD Chips; however, this was not observed in Healthy Chips (**Fig. 3D, E and Extended Data. Fig. 10).** Interestingly, exposure to E2, MPA or the pregnancy-associated hormone mixture decreased overall inflammatory cytokine and chemokine production in Healthy Chips whereas levels of these inflammatory molecules increased when IBD Chips were exposed to the same hormones (**Fig. 3F).**

**Figure 3:**
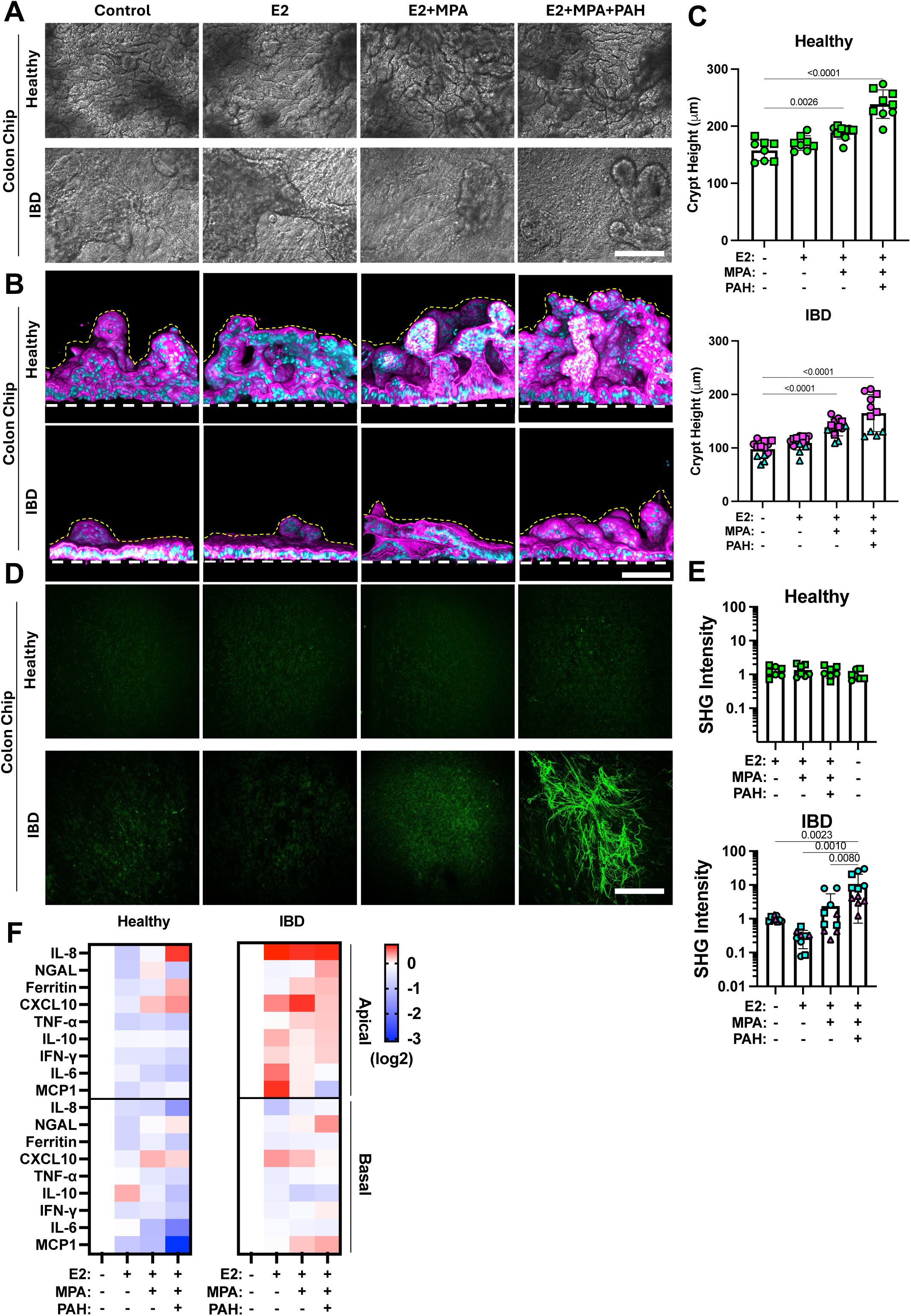
Effects of ovarian hormones in Female Colon Chips. DIC images from above (**A**) and vertical cross-sectional views of confocal microscopic images (**B**) of healthy and IBD Chips 7 days post treatment with exposure to E2, E2 plus MPA, or PAH.. **A**) When viewed from above, exposure to PAH appeared to slightly increase the density of crypt-like structures in both healthy and IBD chips (bar, 500 µm). **B)** When viewed in vertical cross-sections, the number and height of crypt-like structures increased when exposed to PAH as well as E2+MPA in both Healthy and IBD Chips, although the scale of the response was much less in the IBD Chips (bar, 100 µm; magenta: phalloidin, cyan: DAPI; dashed white line, upper surface of chip membrane; dashed yellow line, upper luminal boundary of the crypt). **C)** Quantification of heights of crypt-like structures in Healthy and IBD Chips in the presence or absence of different combinations of the ovarian hormones (E2, MPA, and PAH), as indicated (n=8-12 chips of 2 healthy, 1 CD and 2 UC patients from at least 2 independent experiments). **D)** Second harmonic microscopic images of fibrillar collagen (green) produced by fibroblasts in response to the different hormonal conditions shown in A and B demonstrating a much greater fibrosis response in the IBD Chips compared to Healthy when exposed to pregnancy mimiking hormone combination (E2+MPA+PAH) (bar, 200 μm; n=4-6 chips of 1 CD, 2 UC and 2 Healthy each from 2 independent experiments). **E)** Graphs showing quantification of collagen fibril signal intensity in the stroma of Healthy versus IBD Chips using second harmonic microscopic imaging (numbers indicate *P* values between compared groups, as determined by one-way ANOVA test; n=8-12 chips of 2 healthy, 1 CD and 2 UC patients from at least 2 independent experiments). **F)** Heatmaps showing production of pro-inflammatory cytokine and chemokine proteins in apical (epithelial) and basal (fibroblast) channels of Healthy versus IBD Chips 7 days post treatment with different female hormone exposures, as indicated. Note that exposure to female hormones significantly increased expression of inflammatory factors in both the epithelium and fibroblasts in IBD Chips whereas they suppressed their production in Healthy Chips (numbers indicate *P* values between compared groups, as determined by one-tailed student’s t-test; all data represent mean ± SD). Source data and statistical tests are provided as a Source Data file; *p* values are shown in all figures.

### Stromal Fibroblasts Control Barrier Permeability and Epithelial Inflammation

To better understand the role of fibroblasts in IBD, we took advantage of the synthetic biology nature of human Organ Chip technology to create chips lined by heterotypic tissue recombinants of healthy epithelium with IBD stroma and vice versa. As expected, the homotypic cultures of IBD epithelium and stroma had a compromised barrier with higher permeability than healthy epithelium and stroma recombinants (**Fig. 4A**). However, we discovered that the presence of the IBD fibroblasts was the key driver of barrier disruption as permeability increased significantly when they were combined healthy epithelium in tissue recombinant chips, even though there was no difference in permeability when the different epithelia were cultured alone (**Fig. 4A)**. Interestingly, the permeability of the IBD epithelium alone (i.e., without IBD stroma) was similar to that of healthy epithelium with or without healthy fibroblasts, and tissue recombinants combining healthy fibroblasts with IBD epithelium also did not further alter barrier function, again emphasizing the key role of that the IBD stroma plays in terms of compromising intestinal barrier function.

**Figure 4:**
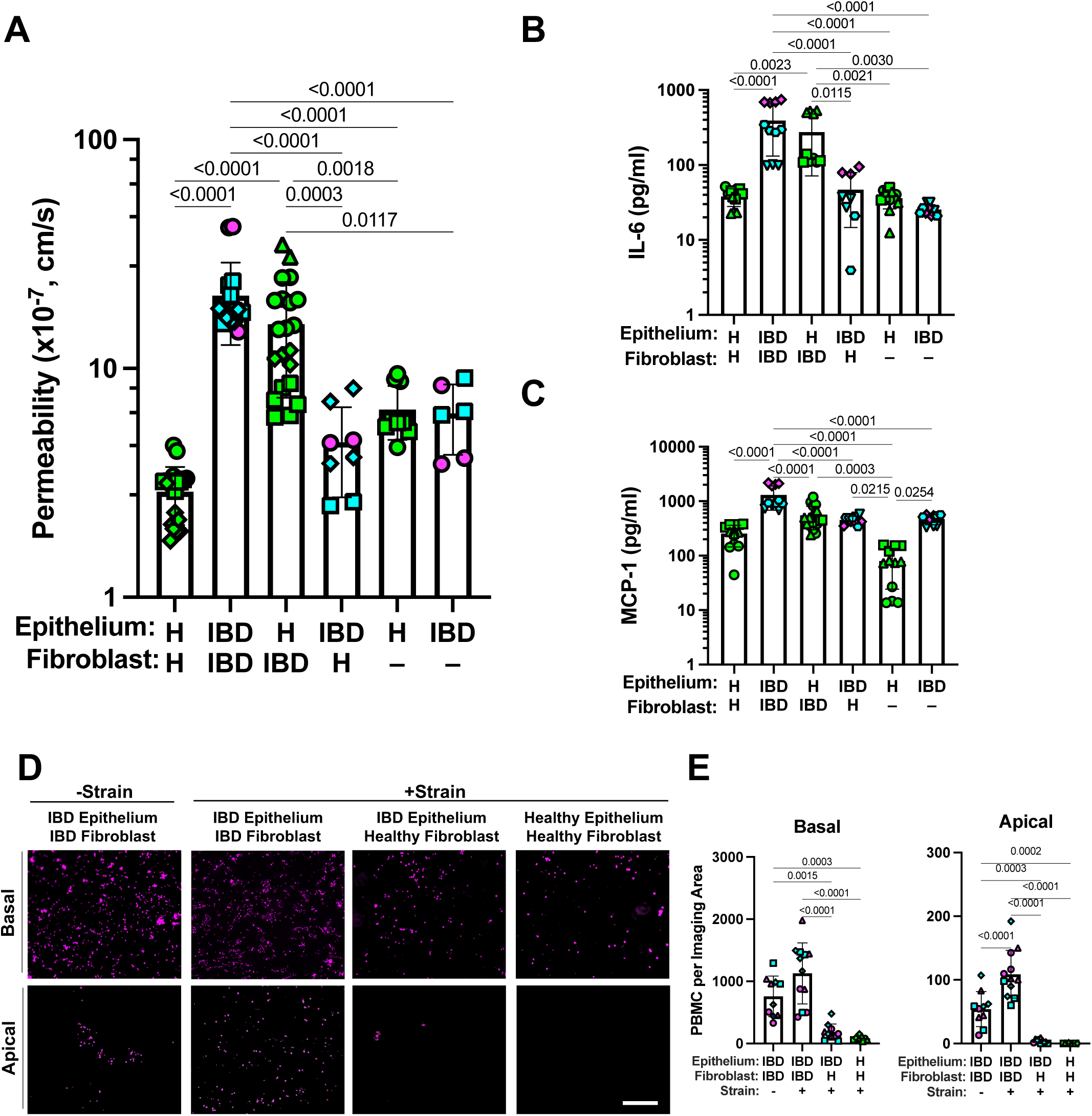
Tissue recombinant chips reveal that the IBD fibroblasts drive compromise of the permeability barrier and inflammation in the epithelium. **A)** Graphs showing that intestinal barrier permeability to cascade blue is much higher in homotypic IBD Chips compared to Healthy Chips and that tissue recombinants of IBD fibroblasts with healthy (H) epithelium results in a similar high level of barrier compromise on-chip. This is not observed in the IBD or healthy epithelium in the absence of fibroblasts or in IBD epithelium with healthy fibroblasts. Numbers indicate *P* values between compared groups, as determined by one-way ANOVA test (n = 3 healthy, green; 2 CD, magenta; and 2 UC, cyan). **B)** Quantification of cytokine protein levels revealed that combination of IBD fibroblasts with healthy epithelium results in increased production of IL-6 and MCP-1 in Colon Chips, whereas this was not seen in healthy epithelium alone or homotypic Colon Chips. Numbers indicate *P* values between compared groups, as determined by one-way ANOVA test (n = 3 healthy, green; 1 CD, magenta; and 2 UC, cyan). **C)** IL-6 and MCP-1 production by IBD epithelium also required the presence of IBD fibroblasts. Numbers indicate *P* values between compared groups, as determined by one-way ANOVA test (n = 3 healthy, green; 1 CD, magenta; and 2 UC, cyan). **D)** Peristalsis-like cyclic strain preferentially stimulates migration of PBMCs (magenta) from the fibroblast stroma in the basal channel (Basal) to the epithelium in the apical channel (Apical) in IBD Chips but not in heterotypic chips with IBD epithelium and healthy fibroblasts or in Healthy Chips. **E)** Quantification of the results from **D** confirmed that IBD Chips exhibited higher baseline recruitment of PBMCs to the surface of the fibroblast stroma in the basal channel and that this was not altered by mechanical strain, whereas IBD epithelium combined with healthy stroma or Healthy Chips only displayed significantly less PBMC recruitment. In contrast, the number of PBMCs that migrated to the epithelium in the Apical channel more than doubled in IBD Chips when they were exposed to peristalsis-like mechanical deformations, and overall migration of PBMCs was minimal when IBD fibroblasts were absent. Numbers indicate *P* values between compared groups, as determined by one-way ANOVA test (n = 2 healthy, green; 1 CD, magenta; and 1 UC, cyan). Source data and statistical tests are provided as a Source Data file.

This controlling effect of IBD fibroblasts on barrier function when combined with healthy epithelium was also accompanied by increased production of multiple inflammatory cytokines and chemokines including IL-6, MCP-1 (**Fig. 4B**), IL-8, CXCL-1, LIF, IL-21, PIGF-1, and MIP-1b (**Extended Data Fig. 11A**) relative to chips lined by healthy fibroblasts and epithelium or healthy epithelium alone. However, the IBD epithelium appeared to constitutively express high levels of all these cytokines regardless of whether or not IBD fibroblasts were present or they were replaced with healthy fibroblasts, except for production of IL-6 and MCP-1 that were further enhanced by the presence of the IBD stroma (**Fig. 4C** and **Extended Data Fig. 11B**). As these two cytokines are known to promote monocyte infiltration into stroma from the vascular system and their differentiation to macrophages cells^32^, we perfused human peripheral blood mononuclear cells (PBMCs) through the basal channel of the Colon Chips. We found that more PBMCs adhered to the stroma and migrated to the epithelial channel in the IBD Chips compared to Healthy Chips (**Fig. 4D**). While application of peristalsis-like deformations did not alter PBMC adhesion to the stroma in the basal channel, it significantly enhanced the migration of these cells to the epithelium in the apical channel, but only in IBD Chips (**Fig. 4E**). However, when we carried out a similar study in a heterotypic tissue recombinant chip containing IBD epithelium with healthy fibroblasts, PBMC recruitment and migration were suppressed and reverted to the low levels observed in Healthy Chips (**Fig. 4D,E**). This finding correlates well with transcriptomic analysis, which revealed that multiple surface receptors for immune cell adhesion are expressed at higher levels in fibroblasts and epithelium in IBD Chips versus Healthy Chips (**Extended Data Fig. 11C**). IBD Chips co-cultured with PBMCs also showed increased production of IL-6 and IL-8 in both their apical and basal channels, whereas this was not observed when Healthy Chips (**Extended Data Fig. 11D**).

### Modeling Early Cancer Progression in Human IBD Chips

Patients with IBD have a higher incidence of colorectal cancer formation^33^ and this also has been demonstrated in animal IBD models by exposing them to the carcinogen ENU^34^. When we exposed Healthy and IBD Chips created with cells from multiple different donors to ENU (10 μg/ml) for 3 weeks, we observed a decrease in the height of crypt-like structures (**Fig. 5A,B**) and an increase in barrier permeability (**Fig. 5C**) in both in healthy and IBD Chips. However, ENU had a much greater impact on inflammatory cytokine secretion in the IBD Chips (**Fig. 5D**) and it resulted in a greater reduction of E-cadherin expression in IBD Chips versus Healthy Chips, which was accompanied by a concomitant increase in *β*-catenin translocation to the nucleus (**Fig. 5E,F**). Similar activation of the *β*-catenin signaling pathway has previously been shown to be associated with colorectal cancer progression^35^. In addition, ENU treatment promoted enlargement of epithelial nuclei and a decrease in their roundness, which are also known to accompany early carcinogenesis^36^. Importantly, shallow pass genome sequencing revealed increased gene mutations and duplication in IBD Chips, but not in Healthy Chips (**Fig. 5G**). Interestingly, the CD chips exhibited a higher level of gene amplification than chips lined with cells from UC patients, and an existing mutation in one CD patient was further amplified after ENU exposure.

**Figure 5:**
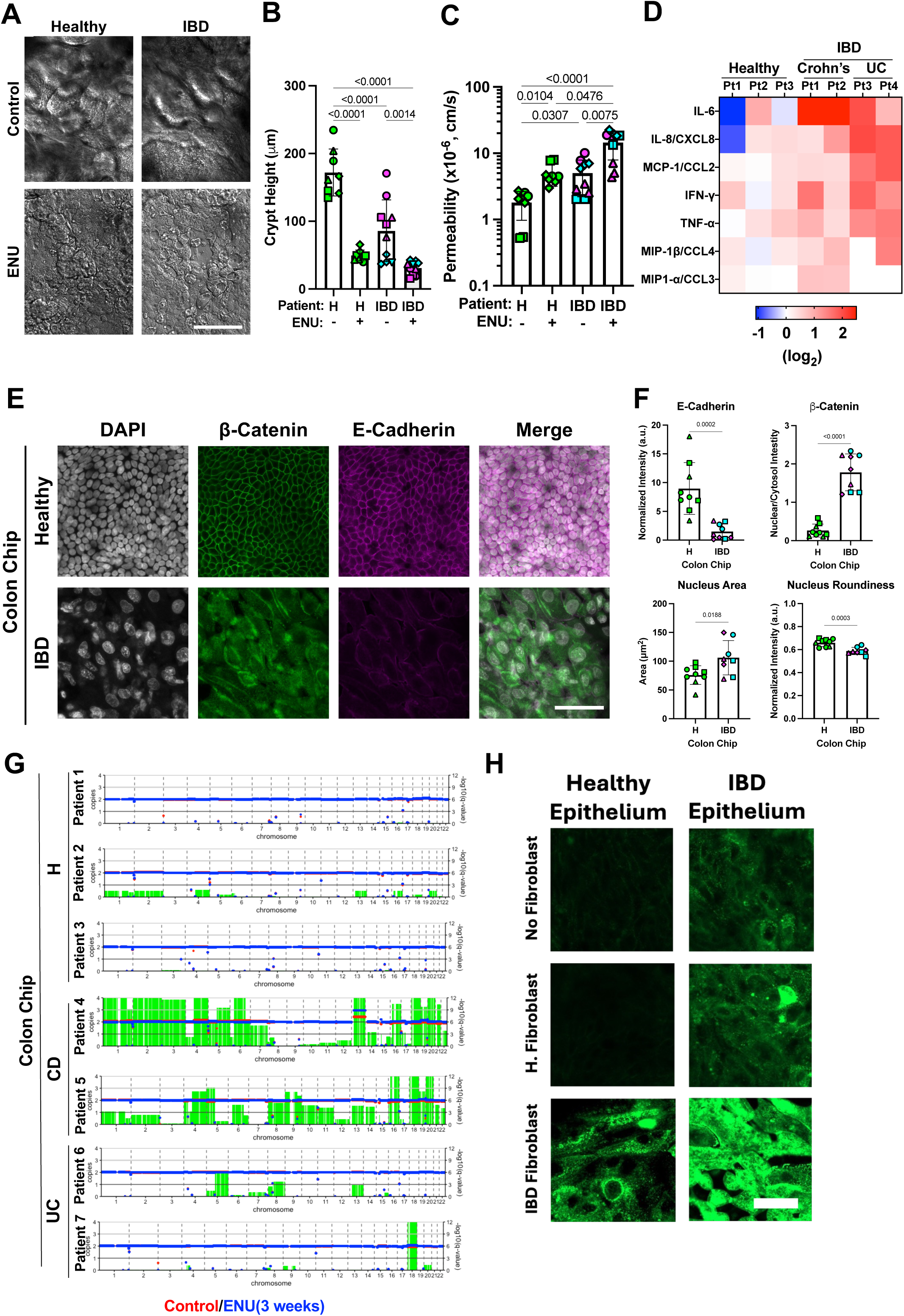
Exposure to carcinogen preferentially promotes mutagenesis and early steps in carcinogenesis in IBD Chips. **A)** DIC images of the epithelium when viewed from above in Healthy (H) and IBD Chips after 21 days of culture in the presence or absence of the carcinogen (ENU) showing that undulating crypt-like structures are lost in both Healthy and IBD Chips after exposure to ENU (bar, 500 µm). **B)** Quantification of results from **A** showing that ENU decreases height of the epithelium in both healthy and IBD Colon Chips. Numbers indicate *P* values between compared groups, as determined by one-way ANOVA test (n = 3 healthy, green; 2 CD, magenta; and 2 UC, cyan). **C)** Epithelial barrier permeability increased as a result of exposure to ENU in Healthy and IBD Chips. Numbers indicate *P* values between compared groups, as determined by one-way ANOVA test. (n = 3 healthy, green; 2 CD, magenta; and 2 UC, cyan). **D)** Heatmaps showing inflammatory cytokine protein levels measured in outflows from the apical channel of Healthy and IBD Chips 3 weeks post-ENU exposure. Importantly, ENU exposure significantly increased inflammatory cytokine production in IBD Chips made with cells from both CD and UC patients compared to healthy patients. **E)** Confocal microscopic top-view images showing β-catenin and E-cadherin localization and nuclear staining (DAPI) in the epithelium within Healthy and IBD Chips 3 weeks post-ENU exposure (bar, 20 μm). **F)** Quantification of protein staining intensity and morphology reveal that ENU exposure decreases E-cadherin levels and increases nuclear localization of β-catenin in IBD patient-derived Colon Chips compared to Healthy Chips. ENU exposure also resulted in greater nucleus size and less roundness. Numbers indicate *P* values between compared groups, as determined by Student’s t-test (n = 3 healthy, green; 2 CD, magenta; and 2 UC, cyan). **G)** Plots showing copy number changes across all human chromosomes in Healthy and IBD Chips pre-(red) and 3 weeks post-ENU exposure (blue). Green bars indicate amplification of gene sequences for the whole chromosome. Each symbol represents an individual Healthy or IBD patient-derived chip. **H)** Confocal microscopic top-view images showing early stage CRC marker, CEACAM5, expression in healthy and IBD epithelium when they are cultured with with or without healthy fibroblasts or IBD fibroblasts and exposed to ENU for 3 weeks. Source data and statistical tests are provided as a Source Data file.

We then investigated whether the fibroblast phenotype influences the propensity for cancer formation following ENU exposure in healthy versus IBD epithelium cultured on-chip. CEACAM5 is an early stage colorectal cancer (CRC) marker^37^, which is expressed in only CRC tissues and not in healthy and IBD intestinal epithelium in vivo (**Extended Figure 12A**). When we analyzed its expression in epithelium in our heterotypic and homotypic tissue recombinants on-chip, we found that neither healthy nor IBD epithelium expressed CEACAM5 on-chip under baseline conditions without ENU exposure (**Extended Data Fig. 12B).** Importantly, healthy epithelium exposed to ENU expressed CECAM5, but only when cultured with IBD fibroblasts (**Fig. 5H and Extended Data Fig.12C**). The presence of IBD fibroblasts also enhanced expression of CEACAM5 in IBD epithelium when exposed to ENU, although lower levels of CEACAM5 expression were even observed when IBD epithelium was cultured alone or with healthy fibroblasts. A greater reduction in E-cadherin expression (**Extended Data Fig. 12D,E**), along with β-catenin translocation to the nucleus also were seen in IBD epithelium cultured with IBD fibroblasts when exposed to ENU (**Extended Data Fig. 12F**). Taken together, these data show that IBD fibroblasts play a significant role in driving a increased propensity for cancer initiation in intestinal epithelium.

## Discussion

Taken together, these findings show that human Colon Chips created with primary patient-derived colonic epithelium and fibroblasts from both healthy and IBD patients recapitulate many features of their respective healthy and diseased phenotypes that are observed in vivo. Based on its synthetic nature, this model enabled exploration of the roles of stromal fibroblasts, epithelium, and immune cells, as well as fluid flow and peristalsis-like mechanical deformations in the etiology of this disease. Importantly, our results revealed that the stroma is a key driver of the IBD phenotype as it is appears to drive effects on intestinal permeability, inflammation, and immune cell infiltration in human intestinal tissues. The finding that IBD stromal fibroblasts respond differently to physiological peristalsis motions than healthy fibroblasts and exhibits enhanced inflammation and fibrotic responses while fluid flow was maintained constant also suggests that physiological mechanical deformations of intestine during early stages of IBD development may feed back to further enhance inflammation and fibrosis thereby accelerating disease progression. In addition, we found that ovarian hormones influence the IBD phenotype directly by targeting the intestinal stroma and enhancing inflammation and fibrosis..

Finally, we demonstrated that the human IBD Chips also exhibit an enhanced sensitivity to a carcinogen and undergo changes that underlie early stages of carcinogenesis in vitro, which is again consistent with the finding that IBD patients have an increased propensity to develop colorectal cancers. Our finding that intestinal stromal fibroblasts influence immune cell trafficking is consistent with a past study in which knock out of genes expressed in an inflammatory subset of fibroblasts produced a similar effect in a mice DSS colitis model^12^.

These results raise the possibility that human Colon Chips could be used as a preclinical model with IBD patient cells to study the effects of emerging immune cell targeting therapies (e.g., anti-integrin α4β7 to suppress infiltration of circulating T-cells) on IBD stroma versus epithelium and to determine their potential efficacy in a patient-specific manner^39^. In addition, this work might serve as inspiration to develop other stroma-targeted therapies given our findings that the stroma is a key driver of the IBD state in both females and males. The stroma also plays a critical role in intestinal homeostasis by supporting the intestinal stem cell niche growth and modulating immune responses, as well as protection against microbiota^40^.

Another novel feature of this study is its relevance for Women’s Health. Female IBD patients can experienced loose and increased stools during the menstrual cycle as well as greater fetal complications during pregnancy ^31,41^. However, it has not been possible in the past to explore whether or not these symptoms are the result of direct effects of female hormones on intestinal tissues. This is because female hormone research is challenging using animals. For example, mice that are often used for women’s health research^42^ do not menstruate like humans do (e.g., they undergo an estrous cycle) and their immune responses different significantly from human^43^. Because biopsy collection during menstruation or pregnancy is not easily done in a clinical setting, it is also challenging to understand the biological basis for clinical symptoms observed in female IBD patients. Another advantage of our model is that we can measure the direct effects of these hormones on healthy versus IBD intestinal tissues. Our finding that pregancy hormones directly enhance inflammation and fibrosis in Colon Chips, particularly in female IBD Chips could be at least in part explain the exacerbation of IBD symptoms symptoms observed during pregnancy in 30-35% of female IBD patients^38^. We also observed an increase in the height of crypt-like structures in Colon Chips as a result of exposure to pregnancy-associated hormones both in healthy and IBD chips. A similar increase in villi height was observed in the small intestine of healthy pregnant mice, and this increased surface area is thought to support increased nutrient absorption for the fetus.^44,45^ However, our results showing that the colonic epithelium undergo similar expansion in response to pregnancy hormones in both healthy and IBD tissues suggests that this may not be limited to nutrient absorption, and also include water and electrolyte reabsorption. Thus, these data provide new insights into how female hormones influence intestinal physiology in both healthy and IBD patients.

In addition to increased inflammation and decreased mucus production, lack of intestinal motion due to fibrosis is a hallmark of IBD; however, it has not been previously possible to explore whether peristalsis plays a role in earlier stages of IBD progression. We found that peristalsis-like mechanical deformations increased production of the protective mucus layer in Healthy Chips, but less so in IBD Chips, and that these mechanical cues also enhanced inflammation and fibrosis, but only in IBD Chips. This suggests that persistence of physiological peristalsis during early stages of inflammation may promote progression of IBD, and that fibrosis and resultant suppression of peristalsis likely occur at a later stage in this process. This may, in part, occur because the IBD tissues lose their ability to maintain the mucus barrier that protects against dysbiotic microbiome and pathogens that are known to be enriched in the microbiome of IBD patients, such as *Ruminococcus* family members that degrade mucus^46^. This could then lead to more inflammation and a positive feedback loop characterized by enhanced sensitivity to these mechanical deformations resulting in greater increases in inflammation, ECM accumulation, and fibrosis at later stages of the disease, which stiffens the mucosa and physically restricts tissue distension.^41^

Chronic inflammation in patients with IBD is known to be a risk factor for increased cancer formation. Importantly, our results show that human IBD tissues do indeed exhibit an enhanced sensitivity to a carcinogen (ENU) compared to Healthy colon tissues on-chip, as indicated by crypt blunting, loss of adherent junctions, overexpression of the early stage CRC marker CEACAM5, and localization of **β**-catenin to the nucleus as seen in IBD patient tissues that progress to colorectal cancer^47^. Importantly, we also observed a greatly increased inflammatory response as well as accelerated development of cancer-associated mutations and chromosomal amplifications in IBD Chips but not Healthy Chips, even though immune cells were not present. While similar effects have been observed after exposure to ENU in a murine intestine model ^48^, this to our knowledge is the first demonstration of early cancer progression in a preclinical *human* IBD model. Although it has been believed that both CD and UC patients carry a similarly high risk of cancer progression ^49^, many CD patients undergo colectomy early in the course of the disease to alleviate persistent symptoms^50^, and this effectively eliminates their risk of developing colorectal cancer.^59^ Interestingly, although our sampling was small, we found that ENU induced greater gene and chromosomal aberrations in CD patient tissues than in UC patient chips, suggesting that they might exhibit a greater susceptibility to carcinogens. More importantly, use of Organ Chip technology enabled us to create tissue recombinant models, which revealed that IBD fibroblasts play a central role in driving this increased propensity for cancer formation. This result is consistent with finding that cancer-associated fibroblasts also promote a dysplastic transition in metaplastic gastric epithelial cells.^51^ This is a finding which should be followed up in clinical studies in the future.

The incidence of IBD is anticipated to increase in the upcoming years. Limitations in the ability of in vivo models to represent human IBD physiology are evident, as the most commonly used anti-TNF and anti-IL-12/23p40 antibodies, which benefit patients with IBD, are generally not effective in chemically-induced colitis models^52^. The need for modeling IBD using Organ Chip technology also has been acknowledged by leading IBD organizations, such as the Crohn’s and Colitis Foundation^53^. Therefore, we believe this novel microfluidic disease model brings important opportunities for IBD treatment, management, and drug development. As we can analyze outflows from the epithelial and stromal compartments separately and through use of tissue recombinants, this method also may facilitate discovery of potential stool and blood biomarkers of IBD progression and cancer risk as well as response to therapy. Use of tissue recombinants is a powerful method for analyzing epithelial-mesenchymal interactions in commonly used developmental biology research, which as we show here also may be applied to confront similar questions relating to human epithelial-stromal interactions, which are challenging to address in vivo. In addition to enabling study of the effects of female hormones on IBD, and it also could allow investigation of other IBD subcategories, including very early onset (VEO-IBD), adolescent IBD, and genetic forms of the disease by lining chips with relevant patient cells.

While this model recapitulates IBD-associated features observed in the human colon, other intestinal regions such as the ileum, duodenum, jejunum, and rectum, could also be modeled using IBD tissues collected from these sites. Modeling these additional regions, particularly the small intestine, would be valuable for investigating absorption and metabolism-related complications in IBD. Notably, Organ Chip technology has already enabled the modeling of these regions using tissues collected from healthy patients^54–56^. Diet can also influence exacerbation of colitis and a small intestine IBD model could be beneficial to investigate diet-associated features in IBD, given that short chain fatty acids (SCFAs), such as butyrate, produced by healthy microbiome can affect IBD remission^57,58^. While we modeled epithelial-stromal interactions in IBD chips, other IBD specific features. such as ileitis-associated fat creeping. can be modeled as well by including adipocytes in the model. In this study, we co-cultured immune cells alongside disease-specific epithelial cells and fibroblasts. This capability can enable evaluation of immune cell-targeting therapeutics, such as α4β7, TL1A, JAK, TNF-α and IL-23 inhibitors^59^, alone or when combined with each other for their potential downstream effects on mucosal tissues and associated complications. Particularly fibrosis has been an unmet challenge in management of IBD in patients as well as other fibrotic diseases (e.g. cystic fibrosis, non-alcoholic fatty liver disease, nonalcoholic steatohepatitis and idiopathic pulmonary fibrosis). Finally, given the synthetic nature of the model and as living gut microbiome can be co-cultured in these Intestine Chips for extended times^54,60^, this experimental system also may be used to study how other environmental factors (such as pathogens) contribute to development of the IBD phenotype in the future.

## Methods

### Acquisition of human colon tissue and cells

Healthy colon tissue samples were collected from macroscopically grossly unaffected regions of the colon of de-identified healthy adult patients undergoing endoscopy for non-IBD related complaints, such as diverticulitis. IBD tissue samples were collected from tissue resections from IBD patients with either CD disease or Ulcerative Colitis. Informed consent and developmentally appropriate assent were obtained at Massachusetts General Hospital and McGill University Health Centre from the donors. All methods were performed in accordance with the Institutional Review Board of Massachusetts General Hospital approval (protocol number IRB-2015P001859) and McGill University Health Centre (protocol number REB-2007-856). We generated epithelium-derived organoids (colonoids) that were cultured in Matrigel using published methods^13,54^ and isolated stromal fibroblasts from the same IBD tissues from 3 CD and 4 UC patients, which were cultured in fibroblast medium (FM, CC-3132, Lonza, MA) in 75 cm^2^ flasks (3531135, Falcon, Durham, NC) with Primocin (100 μg/ml) replaing the GA supplement. Healthy colonoids and fibroblasts were isolated from areas with normal histology adjacent to regions of diverticulitis from 4 otherwise healthy patients (**Supplementary Table 1**). The donor cells used across the experiments were provided in **Supplementary Table 2**. All reagents and resources were provided in **Supplementary Table 3**.

### Human Colon Chip Cultures

Our methods for creating and culturing human Colon Chips lined by primary intestinal organoid-derived epithelium using commercially available Organ Chip Devices (Basic Research Kit; Emulate Inc.) have been described previously^13,61^ and are described in greater detail in Supplementary Materials. Colonoids cultured were released from Matrigel and fragmented by treatment with TrypLE Express Enzyme (12605010; Thermo Fisher Scientific), before being seeded in the apical channel of the 2-channel Organ Chip devices and incubated overnight at 37° C under 5% CO_2_ to promote adhesion. One day later, channels were washed with Expansion Medium (EM) to remove unattached cells, and then perfused with EM (60 µL/hr). At this time, the cultured fibroblasts were detached enzymatically and seeded in EM in the basal channel for 2 h under static conditions to promote adhesion. After washing, EM was then perfused through both channels at 60 μL/h. After 3 days of co-culture during which both epithelial and fibroblast monolayers were established, EM perfused through the apical channel was changed to Hank’s balanced salt solution (HBSS) with calcium and magnesium (21-023-cv; Corning) supplemented with 100 mg/mL Primocin (ant-pm-1; InvivoGen) while EM remained flowing through the basal channel. For experiments exploring the role of physiological peristalsis-like motions, cyclic mechanical strain (10% Strain 0.15 Hz) was applied to the tissues by applying cyclic suction to side chambers of the flexible device^54,61^ for 2 weeks after seeding. Methods for morphological analysis, cytokine quantification, flow cytometry, transcriptional analysis, and atomic force microscopy can be found in **Supplementary Information**.

### Maternal Hormone Treatment

Established Healthy and IBD Colon Chips were exposed to different combinations of hormones from basal channel to recapitulate either the menstrual cycle or first trimester of pregnancy. Chips were treated with 17β-estradiol (E2, 10 nM, Sigma, E4389) for 3 days before being exposed to medroxyprogesterone 17-acetate (MPA, 1 μM, Sigma, 46412-250MG) for an additional 4 days to mimic the menstrual period of the cycle. Chips perfused for 3 days with E2 were then treated with E2, MPA, Prolactin (PRL, 20 ng/mL, VWR, 10770-682), Human Placental Lactogen/CSH1 (hPL, 20 ng/ml, R&D, 5757-PL), and Human Chorionic Gonadotropin (HCG, 1 μg/mL, VWR, IC0219859110) for 4 days to recapitulate the first trimester of pregnancy as described previously^62,63^.

### Immune Cell Recruitment Assay

Colon Chips were cultured for 2 weeks before human peripheral blood mononuclear cells (PBMCs) were added to the cultures for studies on immune cell adhesion and migration. PBMCs were collected from anonymous donors undergoing stem cell mobilization at the Massachusetts General Hospital (MGH) under Institutional Review Board approved protocol #2015P001859. PBMCs were freshly isolated by density centrifugation using the Lymphoprep protocol (StemCell Technologies, #07801), resuspended in 10 ml of DMEM containing 10% FBS(10082-147) and 1% Penicillin-Streptomycin (P4333-100ML), and labeled with 10 µl of CellTracker Deep Red at a final concentration of 10 µM (Invitrogen, #C34565) for 20 minutes at 37°C. The labeled cells were centrifuged and resuspended to a concentration of 5 × 10⁷ cells/ml and 25 µl of the PBMC suspension was added to the lower channel (see **Supplementary Information** for more details).

### Analysis of effects of carcinogen exposure

To explore if the chip models can be used to study the effects of intestinal tissue to carcinogen exposure in vitro, Healthy and IBD Colon Chips were exposed to 10 μg/mL of N-ethyl-N-nitrosourea (ENU, Millipore Sigma, N8509), which was flowed through both the apical and basal channels at 60 μL/hr. Chips were harvested for DNA isolation three weeks after ENU exposure. Briefly, chips were washed with DPBS (-/-) and incubated with TrypLE consisting of 2 mg/mL collagenase-I (Thermo Fisher Scientific, 17100-017) and 10 μM Y-27632 (Y0503; Sigma-Aldrich, St. Louis, MO) at 37°C for 1 h. Single cells were washed with DPBS (-/-) and DNA was isolated by following the manufacturer’s protocol (Qiagen Micro DNA isolation kit, Cat: 56304 Qiagen, Germany). Shallow whole genome sequencing was performed in two batches using different methods (see **Supplementary Information**). The raw sequence reads were mapped to *Homo sapiens* GRCh38 with Burrows-Wheeler Alignment Tool v0.7.17^64^. Reads with a mapping quality of less than 37 were removed from further analysis. The PCR duplicates and base quality recalibration were subsequently carried out using Genome Analysis Toolkit (GATK) v4.4.0^65^ and Picard v2.27.5. The copy number of the genome was then estimated and profiled with ACE v3.16^66^ and QDNAseq v1.35.0 R package^67^ using 100 Mb fixed-sized bins.

### Statistical Analysis

Data in all graphs are expressed as mean ± standard deviation (SD) and significant differences between multiple groups were determined using an unpaired Students t-test, one-way analysis of variance (ANOVA) or two-way ANOVA with Tukey test for correction unless otherwise indicated in the figure legends. P<0.05 was considered significant. Data were analyzed using GraphPad Prism 8 software (GraphPad Software, San Diego, CA).

## Data Availability

Data that support the findings of this study are available within the paper and its Supplementary Information files. The RNA sequencing data have been deposited in the Gene Expression Omnibus (GEO) with the accession code GSE277964. Any other material including the R code used for data analysis is available upon reasonable request.

## Acknowledgement

This work was funded in part by Cancer Research UK (C19767/A27145 to D.E.I.) and by the Wyss Institute for Biologically Inspired Engineering at Harvard University as well as by an NIH training grants (5T32DK007199-44 and 5T32EB016652-10 to A.Ö.) and Wyss Technology Development Fellowship to A.Ö. We thank Harvard Digestive Diseases Center Organoid Core for providing L-WRN conditioned media (P30DK034854).

## Contributions

A.O., S.H. and D.E.I designed the research; A.O., G.E.M, R.R.P. A.S. K. C., E. C., J.P. performed the experiments; A.O., G.E.M., R.R.P. A.S., K.C., E.C. analyzed and interpreted the data; A.O. and G.E.M. established healthy and IBD human organoid and fibroblast biobanks; V. H., M. S. and A.O. performed bioinformatic analysis; D.B.C., L.E.F., R.R., L.B. provided the clinical samples; A.O. and D.E.I. acquired the funding; A.O. wrote the articled with input from G.G., S.H. and D.E.I.; and all authors reviewed, discussed and edited the manuscript.

## Data and code availability

Data that support the findings of this study are available within the paper and its Supplementary Information files. The RNA sequencing data have been deposited in the Gene Expression Omnibus (GEO) with the accession code GSE277964. All data reported in this study are available in the Source Data files. Any other material including the R code used for data analysis is available upon reasonable request.

---

**Extended Data Fig. 1.**
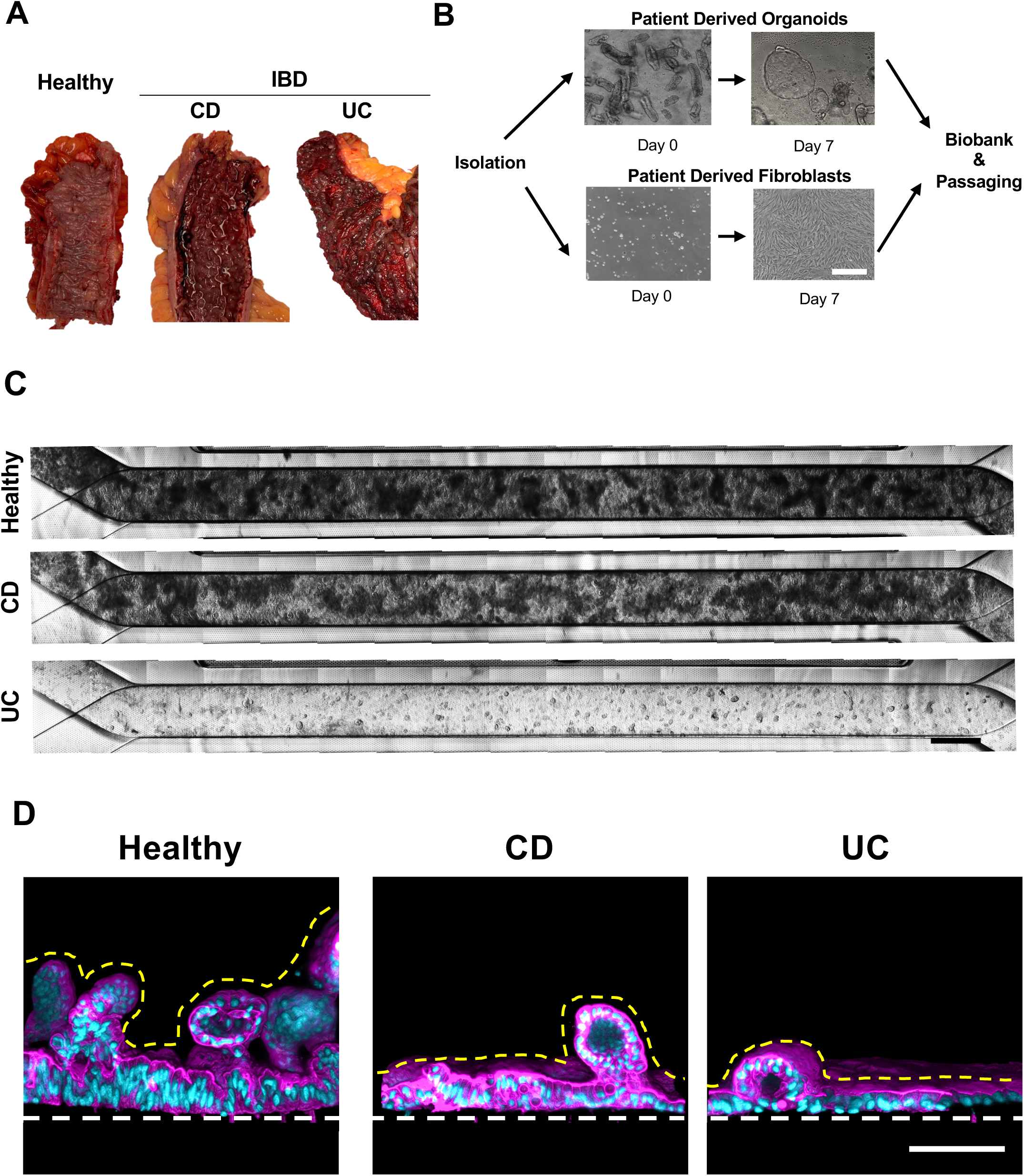

**Extended Data Fig. 2.**
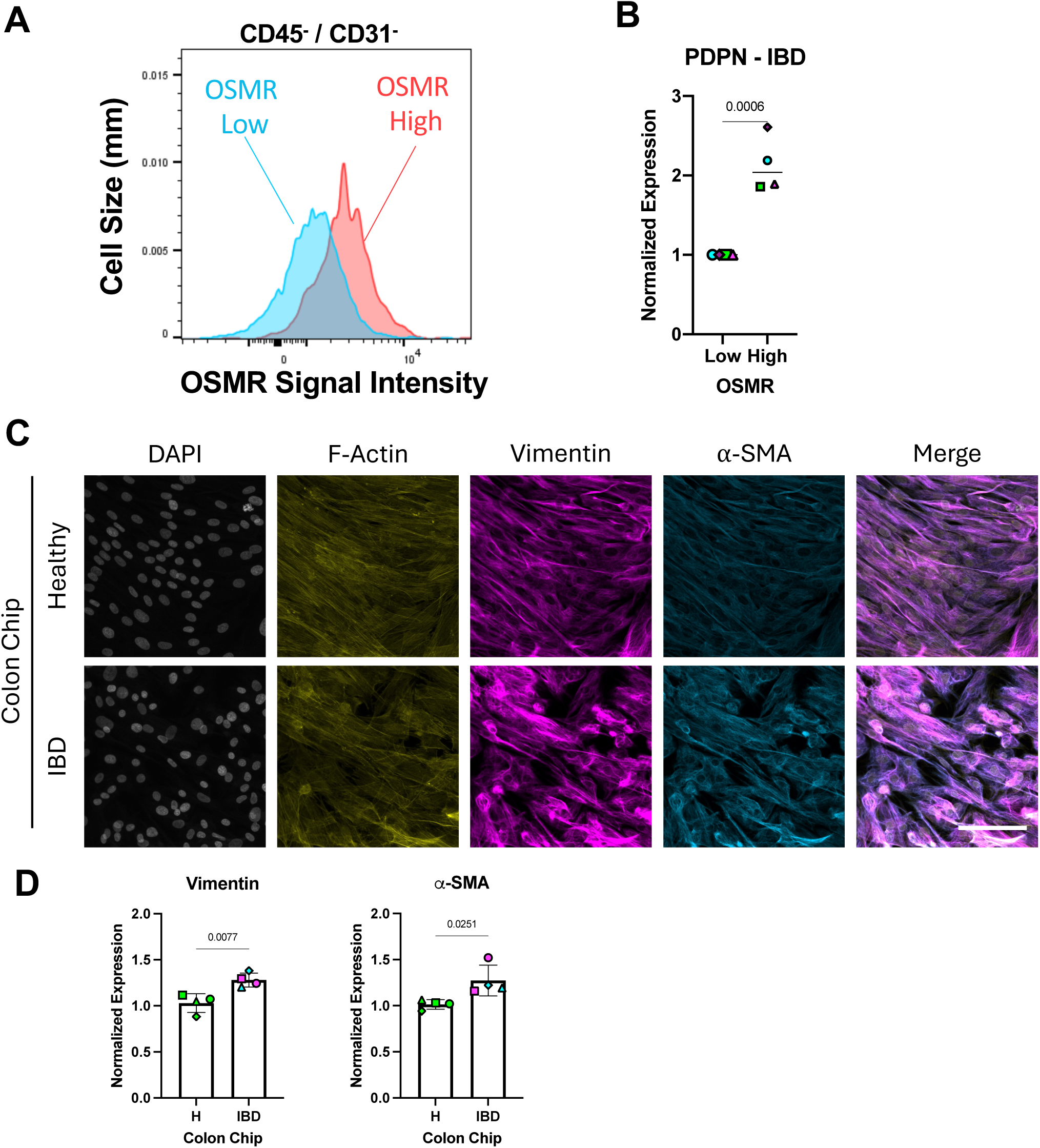

**Extended Data Fig. 3.**
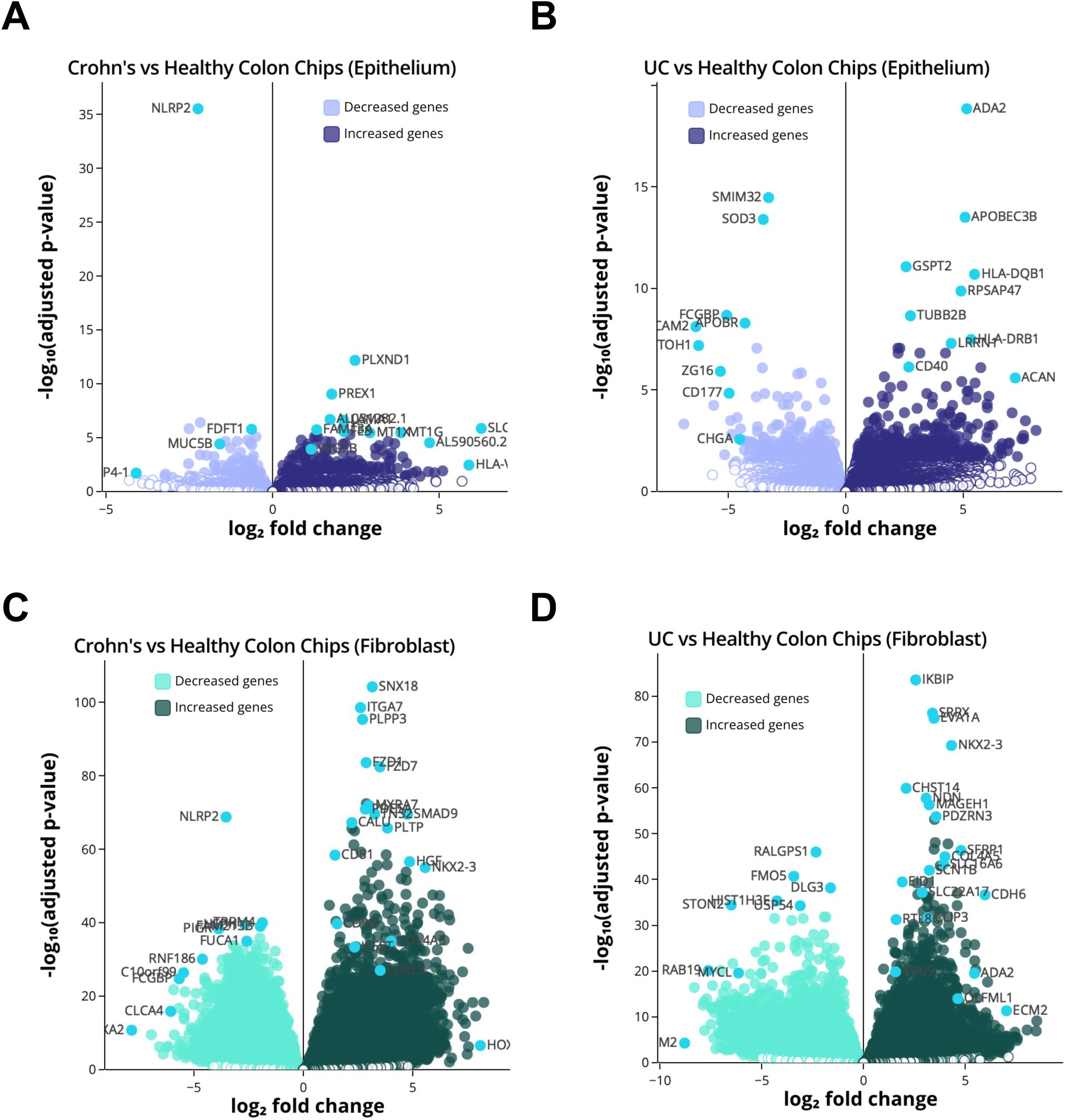

**Extended Data Fig. 4.**
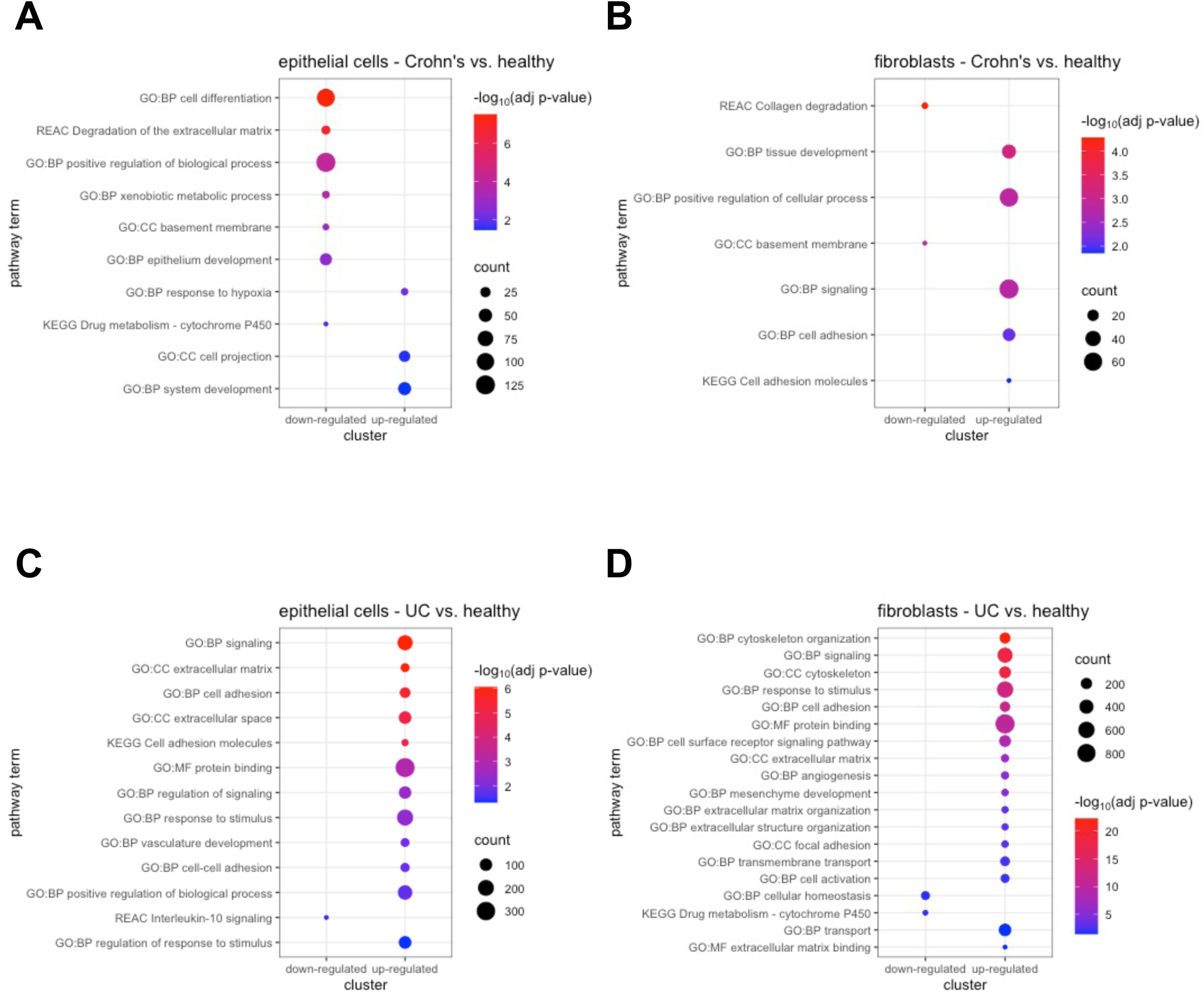

**Extended Data Fig. 5.**
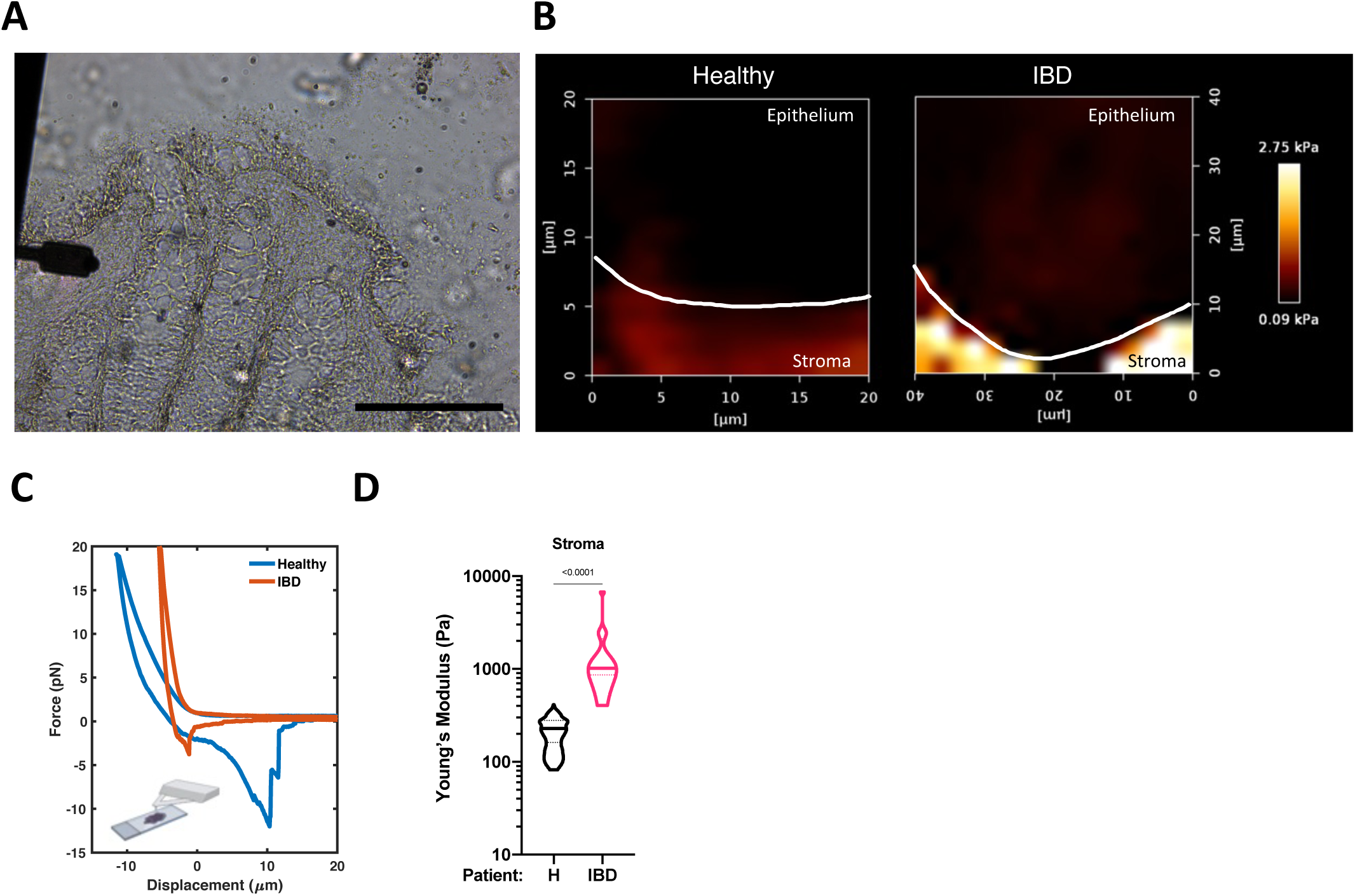

**Extended Data Fig. 6.**
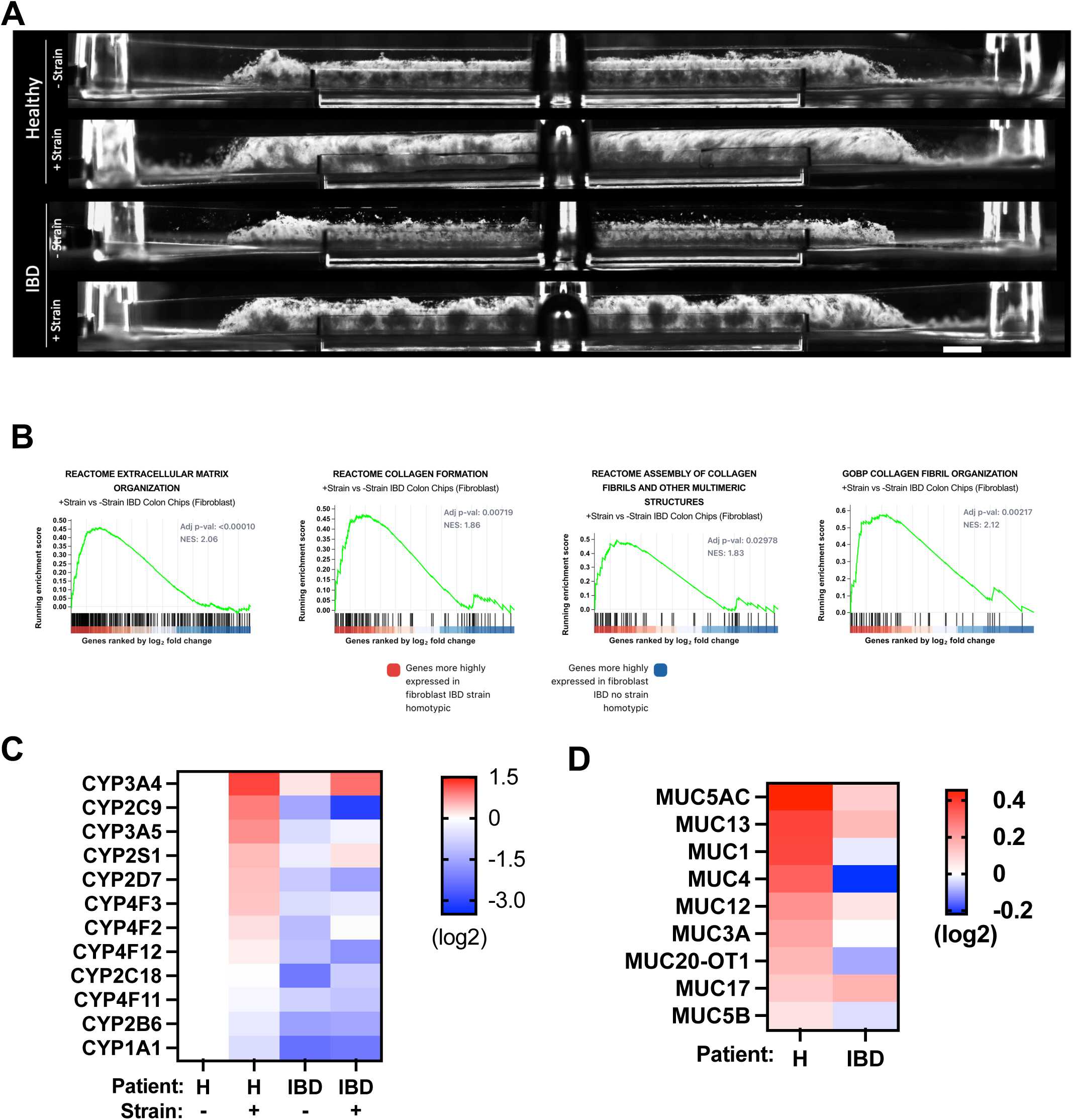

**Extended Data Fig. 7.**
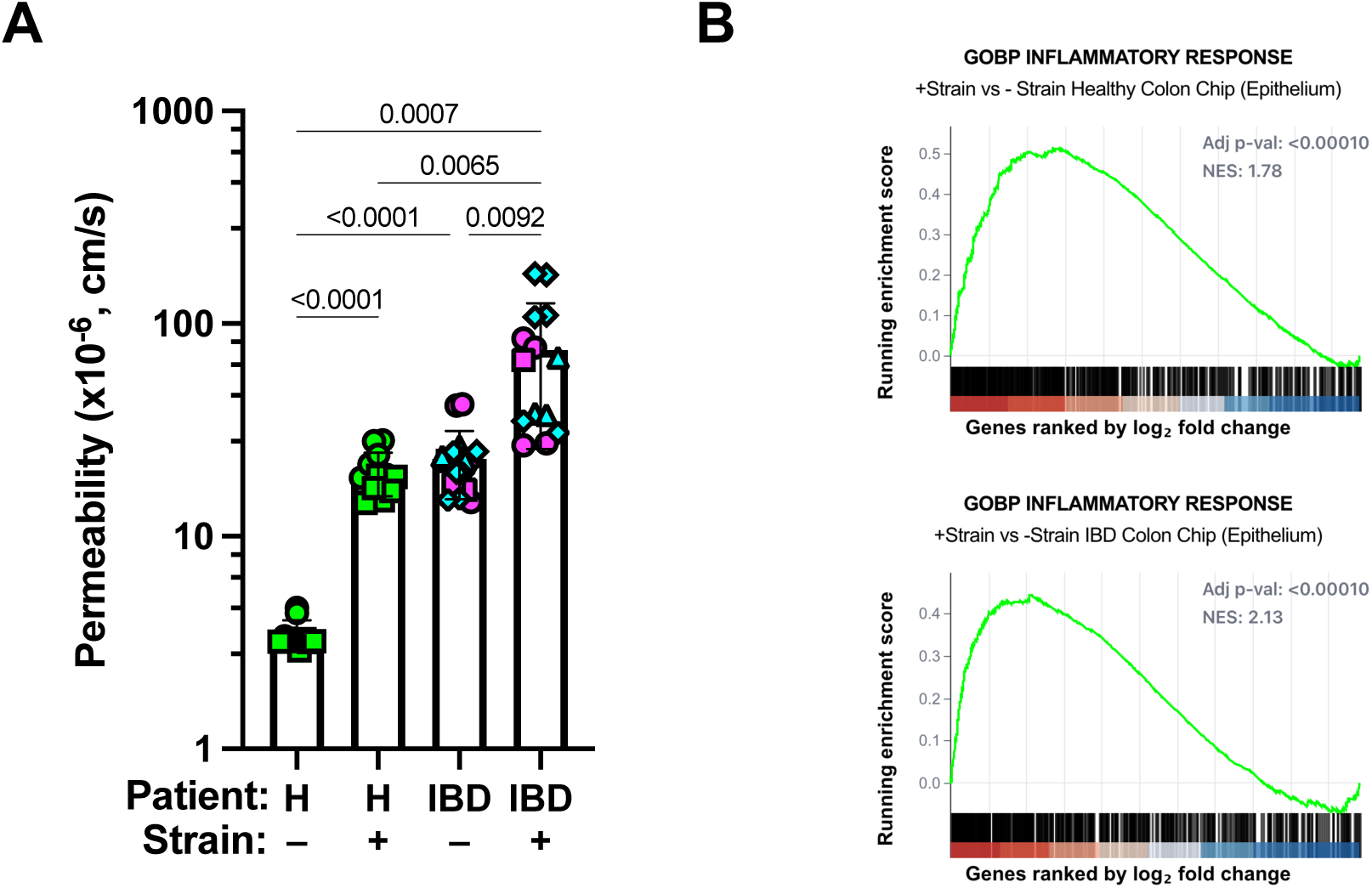

**Extended Data Fig. 8.**
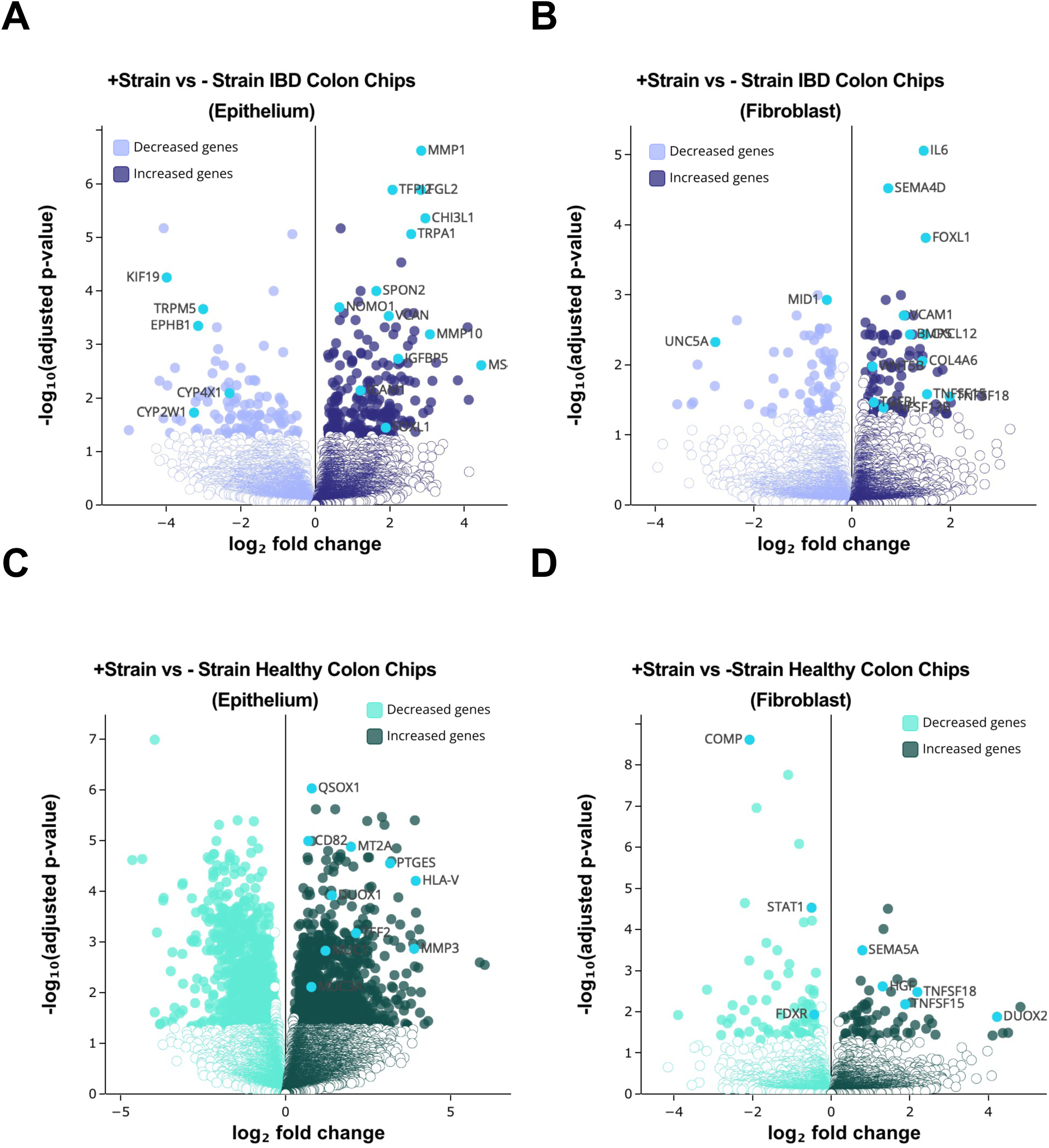

**Extended Data Fig. 9.**
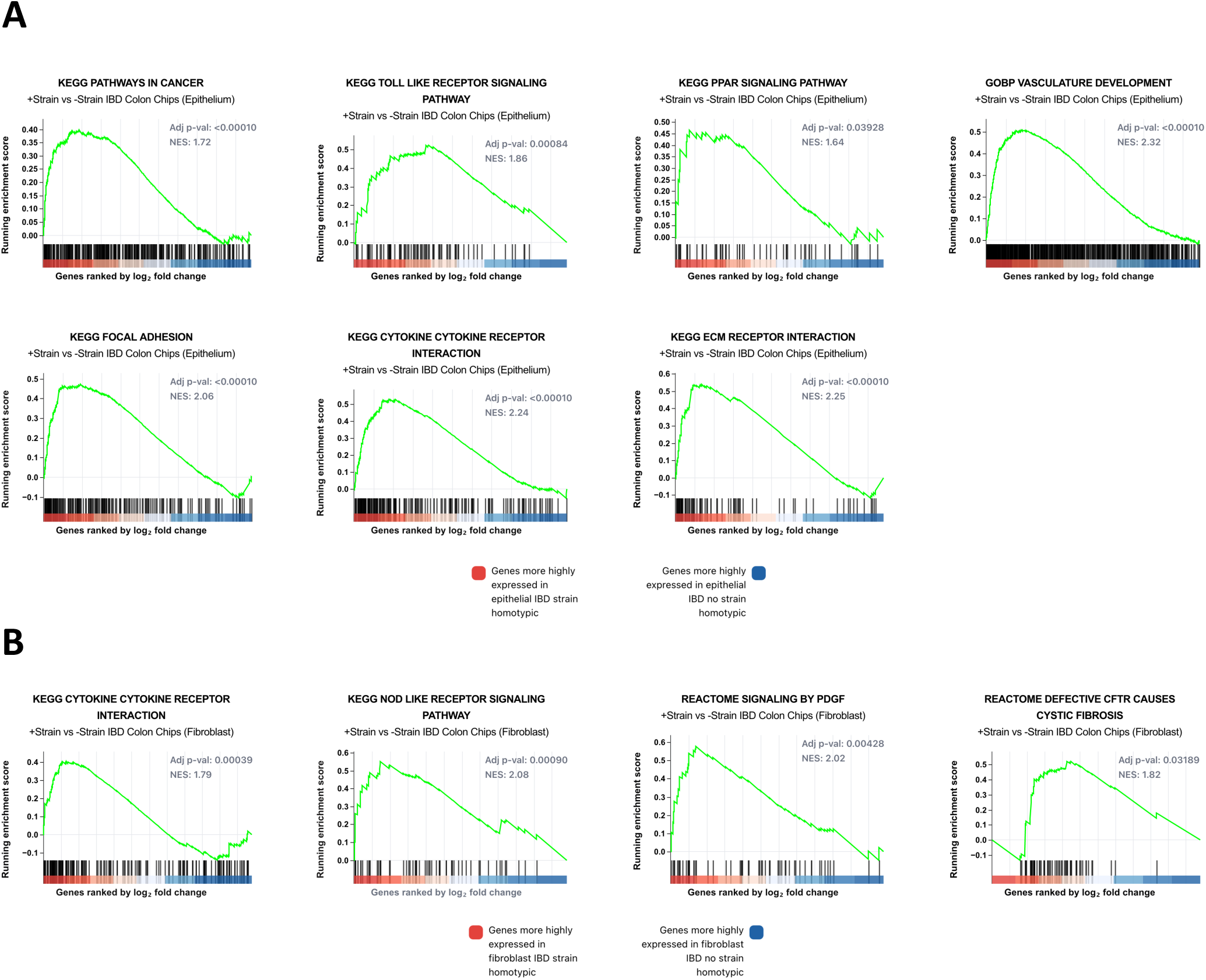

**Extended Data Fig. 10.**
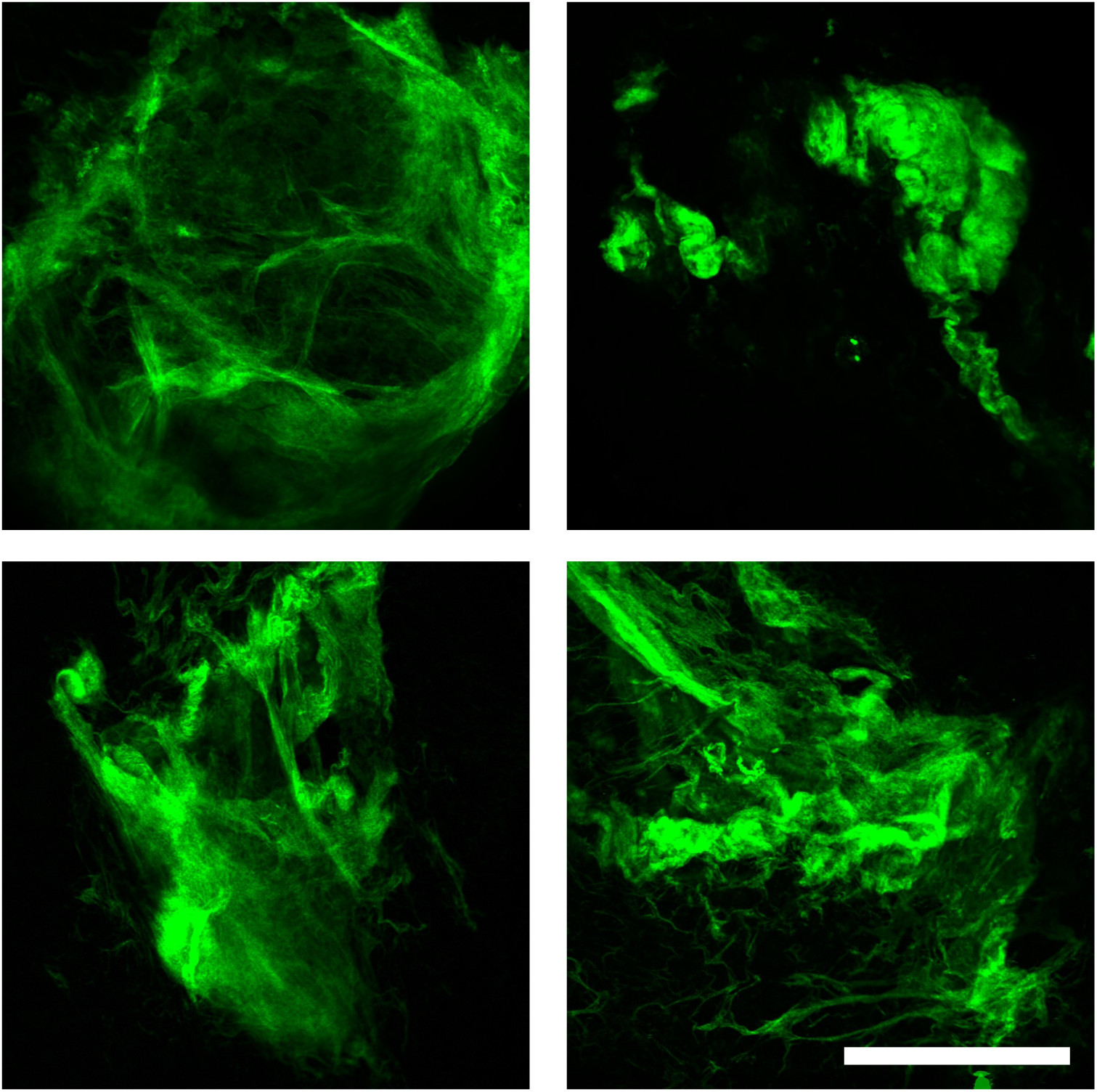

**Extended Data Fig. 11.**
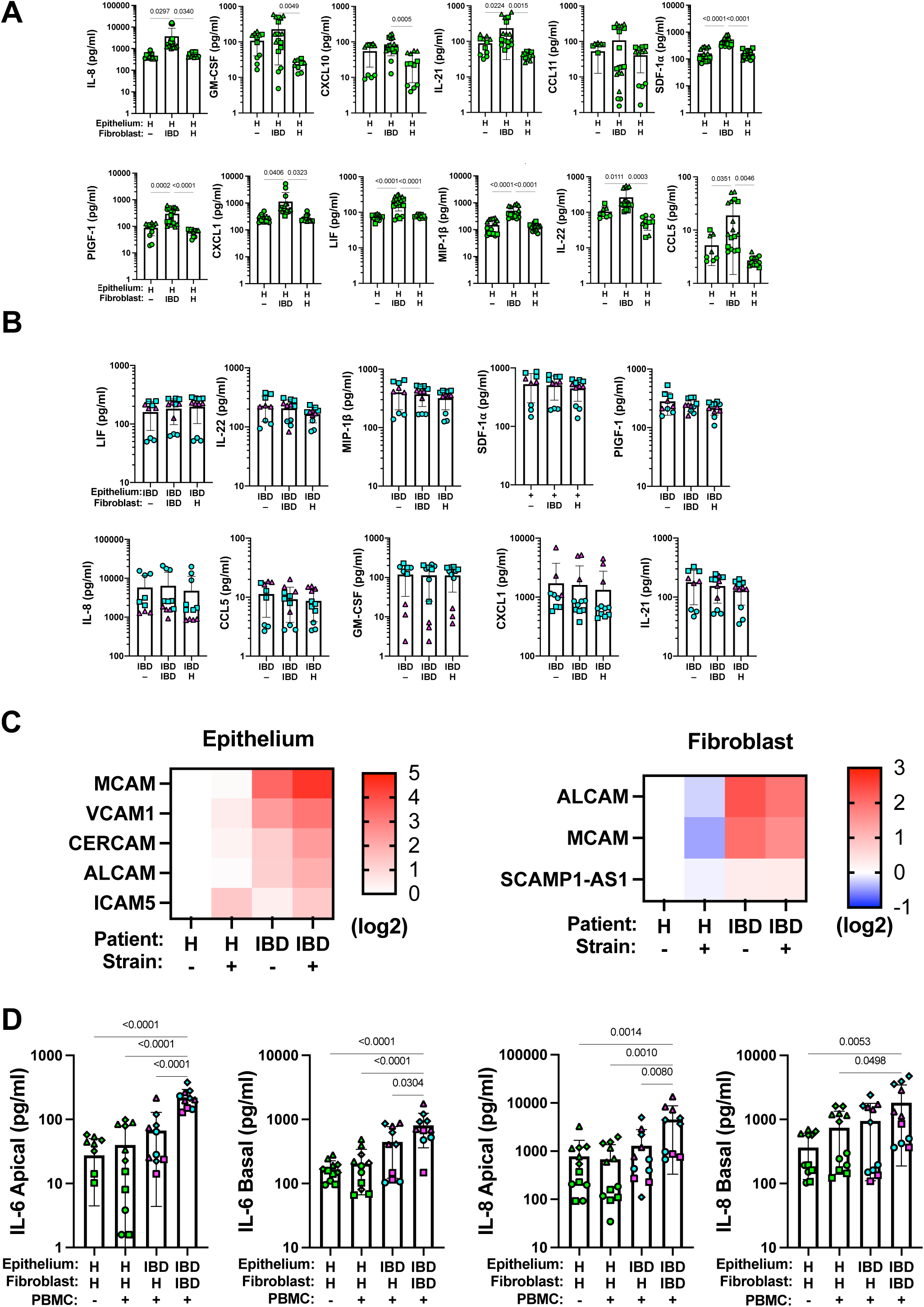

**Extended Data Fig. 12.**
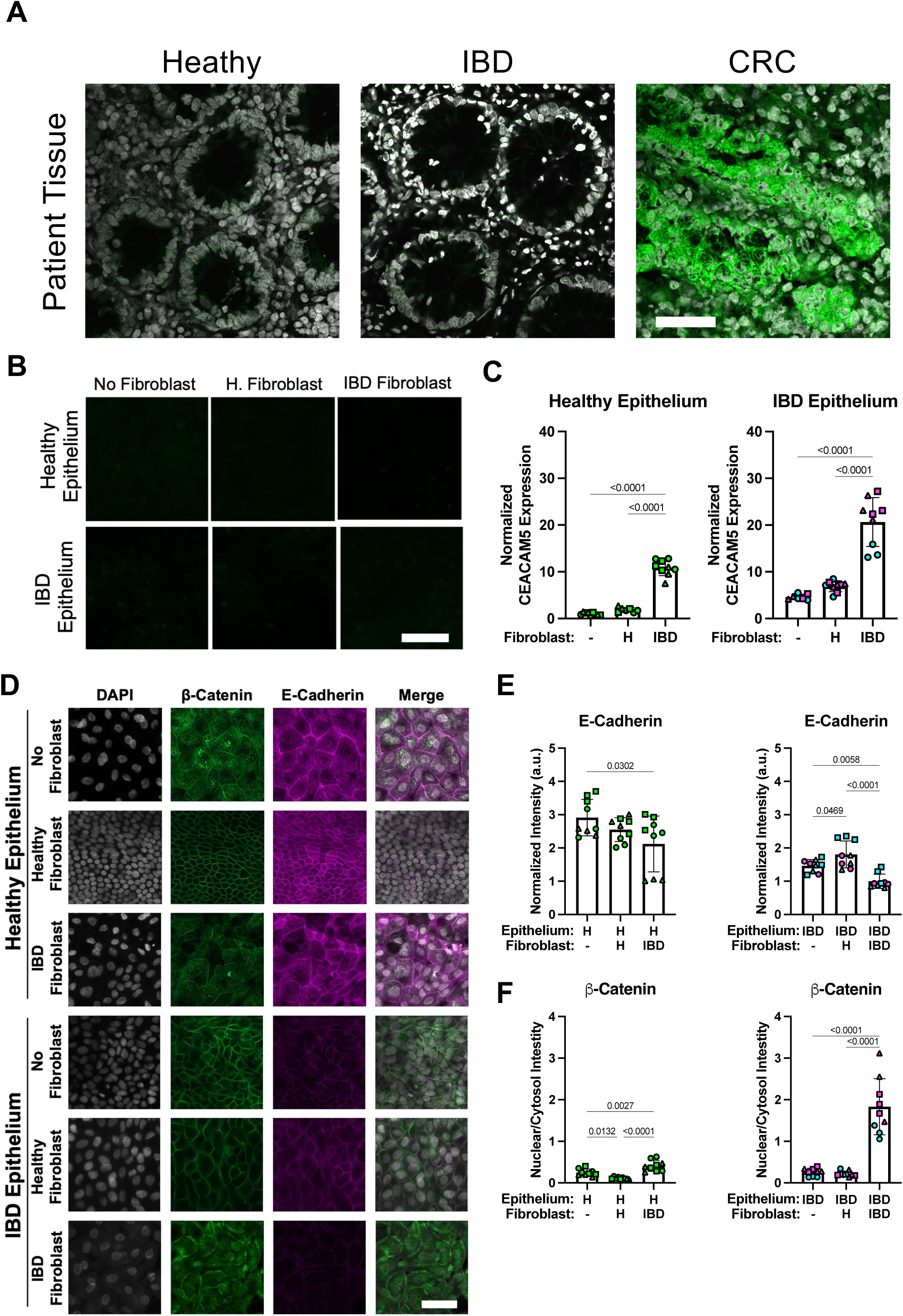

**Supporting Data Fig. 1.**
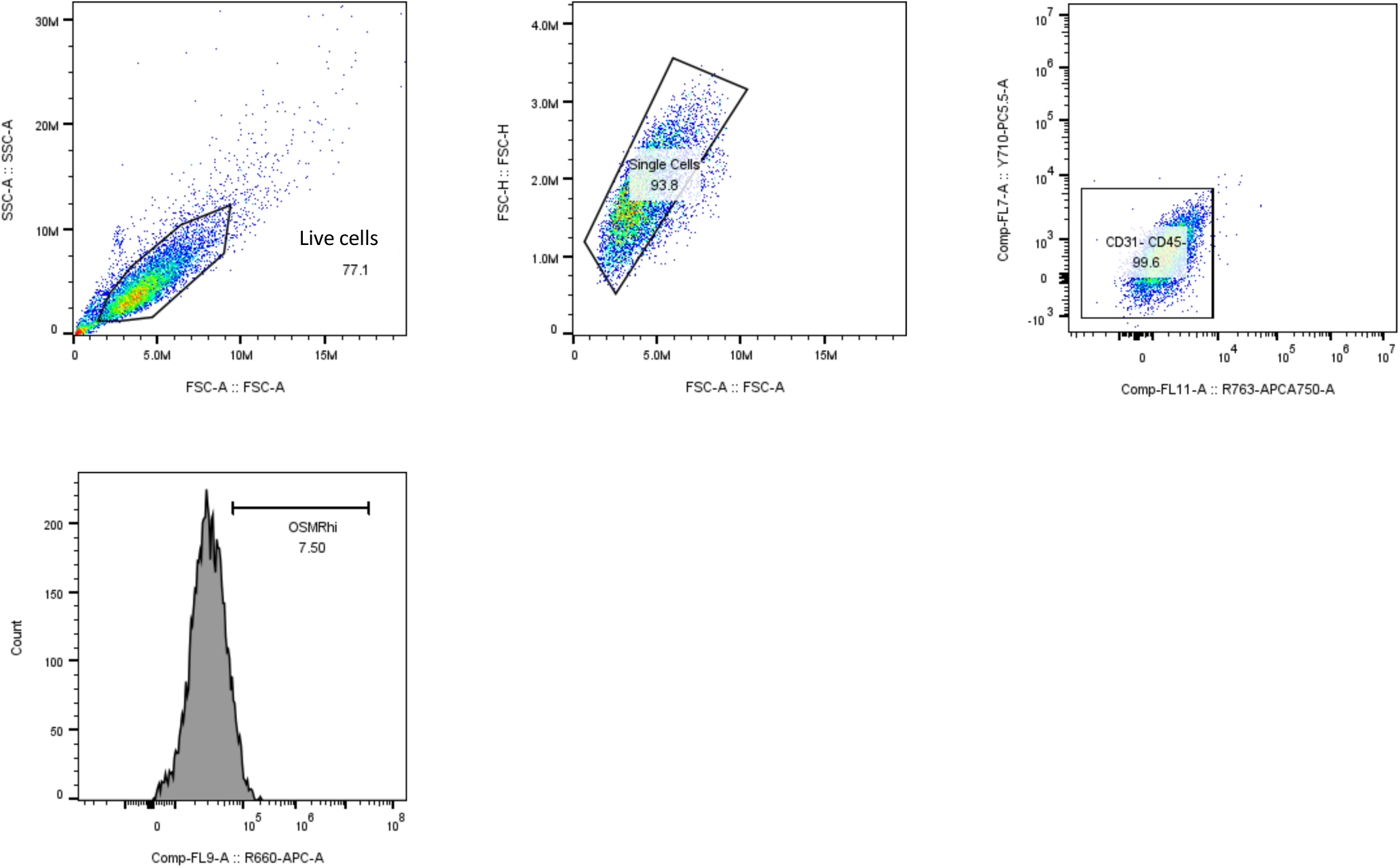

