## Extended Data for "Human Organ Chips Reveal New Inflammatory Bowel Disease Drivers"

### Methods

##### Acquisition and culture of human colon cells

For isolation of human colon cells from patient tissue specimens, colon resections were processed by removing epithelium with lamina propria and digesting the entire specimen with 2 mg/mL collagenase I (17100- 017; Thermo Fisher Scientific, Waltham, MA) supplemented with 10 μmol/L Y-27632 (Y0503; Sigma-Aldrich, St. Louis, MO) for 30 min with occasional agitation. The digested mixture of tissue and cells was divided into 2 tubes, one for colonoid creation and the other for fibroblast isolation. Colonoids were grown embedded in growth factor reduced Matrigel (356231, lot 7317015; Corning, Corning, NY), and the colonoid expansion medium (EM) was composed of Advanced Dulbecco's modified Eagle medium F12 (12634-010; Thermo Fisher Scientific) containing L-Wnt3a, R-spondin, noggin–conditioned medium (65% vol/vol) (produced by the CRL-3276 cell line; American Type Culture Collection, Manassas, VA), 1N GlutaMAX (35050-061; Thermo Fisher Scientific), 10 mmol/L HEPES (15630-106; Thermo Fisher Scientific), recombinant murine epidermal growth factor (50 ng/mL) (315-09; Peprotech, Rocky Hill, NJ), 1N N2 supplement (17502-048; Thermo Fisher Scientific), 1N B27 supplement (12587001; Thermo Fisher Scientific), 10 nmol/L human (Leu15)-gastrin I (G9145; Sigma-Aldrich), 1 mmol/L N-acetyl cysteine (A5099; Sigma- Aldrich), 10 mmol/L nicotinamide (N0636; Sigma-Aldrich), 10 mmol/L SB202190 (S7067; Sigma-Aldrich), 500 nmol/L A83-01 (2939;

Tocris, Bristol, UK), Primocin (100 mg/ mL) (ant-pm-1; InvivoGen, San Diego, CA), which was supplemented with 10 μmol/L Y-27632 (Y0503, Millipore Sigma,MA). Isolated fibroblasts were cultured in Fibroblast Medium (FM) in T-75 flasks (3531135, Falcon, Durham, NC) using fibroblast basal medium (FBM, CC-3131, Lonza) supplemented with fibroblast growth media-2 SingleQuots (CC-4126, Lonza), except GA and supplemented with Primocon (100 μg/ml) which includes insulin, hFGF-B, FBS, and Primocin. Fibroblast biobanks were created at passage 3 and used up to passage 5.

**Human Colon Chip Cultures**

Optically clear Organ Chips made of poly-dimethylsiloxane (PDMS) and containing two parallel microchannels (top, 1 x 1 mm; bottom, 1 x 0.2 mm) separated by 50 μm thick PDMS porous membrane (7 μm pore diameter, 40 μm spacing) were purchased from Emulate Inc (Basic Research Kit). Surfaces were activated using 0.5 mg/mL sulfo-SANPAH solution (A35395; Thermo Fisher Scientific) for 20 min and coated with 0.2 mg/mL collagen type I (354236; Corning) and 1% Matrigel (356231, Corning) in Dulbecco's phosphate-buffered saline (DPBS). During the coating step, colonoids were obtained by dissolving Matrigel using cell recovery solution 354253 (Corning) for 40 min on ice and spun down at 400*g* for 5 min at 4$^{o}$ C. Colonoids were then fragmented with TrypLE Express Enzyme (12605010; Thermo Fisher Scientific), diluted 1:1 in DPBS, and supplemented with 10 μmol/L Y-27632 (2 mL/well of a 24-well plate) for 2 min at 37$^{o}$ C. After adding the same amount of EM with 10 μmol/L Y-27632, colonoids were spun down at 400*g* for 5 min at 4$^{o}$ C and resuspended at 6 x $10^{6}$ cells/mL for seeding.

**Morphological Analysis**

Top-view images of the Colon Chip were acquired using differential interference contrast (DIC) or phase-contrast microscopy (ECHO, San Diego, CA). Fixed samples that were sectioned (200 μm thick) with a vibratome (Leica, VT1000-S) were used for H&E staining to visualize epithelial structures. Side-view dark field imaging was carried out to visualize and quantify mucus formation, as previously described[1]. Briefly, excess PDMS was cut away from each side of the Colon Chips, which were then placed on coverslips with a glycerine solution (11513872; Leica). Images were collected using an Axio Observed Z1 (Zeiss) inverted microscope with 2.5X objective (0.06 NA, 441010-9901; Zeiss) and condenser (0.35 NA, 424241-0000-000; Zeiss) with phase ring 2 in dark field imaging resulting in visualization of fluorescent (white) mucus layer above non-fluorescent cell layer (black). Mucus coverage areas and height were quantified using Fiji software (https:// imagej.net/Fiji).

For immunofluorescence imaging, Colon Chips were fixed with 4% paraformaldehyde (50-980-487) in PBS(+/+) for 30 minutes at room temperature. Chips were either stained directly or sectioned at 200 μm thickness (Leica). Snap-frozen patient samples were sectioned at 10 μm thickness and fixed with 4% paraformaldehyde in PBS at RT for 15 min. All samples were blocked and permeabilized using 0.1% Triton X-100 (Sigma, X100-5ML) and 5% bovine serum albumin (BSA, A9418-50G) in DPBS for 1h at RT. Samples were incubated with primary antibodies in 2% BSA in DPBS overnight. The next day, secondary antibodies were incubated overnight. Finally, samples were counterstained with Hoechst (94403-1ML) for 30 min at RT. Samples were imaged using Zeiss TIRF/LSM 720 confocal microscope with lasers including 352, 488, 561, and 688 coupled with HyD detectors. Acquired images were analyzed either with ImageJ or IMARIS software (Biplane, Zurich, Switzerland). A list of primary and secondary antibodies used is provided in **Table S2**.

The height of extended epithelial crypt-like structures was measured using confocal images of vibratome sectioned healthy and, IBD chips. Crypt-like structure height was quantified by measuring the maximum distance between the upper surface of chip membrane (visualized with dashed white line in the confocal images) and upper luminal boundary of the crypt-like structures (dashed yellow line in the confocal images). ImageJ was used to quantify the distance between the membrane and epithelium.

To visualize ECM accumulation and distribution, fixed Healthy and IBD Colon Chips were analyzed using multiphoton microscopy (Leica SP5 or Stellaris 8) and second harmonic imaging. Apical and basal channels were filled with HBSS before imaging using a 25X (Water) objective, 880 nm wavelength, and a reflectance image was used to visualize fibroblast cell borders. Collected images were analyzed using Image J.

**Cytokine and chemokine analysis**

Apical and/or basal effluents from Colon Chips were collected and analyzed using a custom panel for quantification of multiple cytokines and chemokines associated with epithelial and stromal inflammation, including IL-6, IL-8, MCP-1, SDF-1$\alpha$, PIGF-1, CXCL10, LIF, CCL5, GM-CSF, CXCL-1 IL-22, IFN-$\gamma$, TNF-$\alpha$, MIP-1$\alpha$ and MIP-1$\beta$ (Procartaplex, Invitrogen). Effluent concentrations were determined using a Luminex 100/200 Flexmap3D instrument coupled with the Luminex XPONENT software.

**Flow Cytometry**

After expansion of fibroblasts from healthy, Crohn's, and UC patients, cells were harvested from culture wells using TrypLE, washed by centrifugation, and resuspended in staining buffer composed of 1% FBS (Gibco, 10082-147), 25mM HEPES (Thermo Fisher Scientific, 15630-080), 1mM EDTA (Thermo Fisher Scientific, 15575-020), and 0.05% sodium azide (VWR, BDH7465-2) in DPBS (Gibco, 14190-144). The resulting cell suspension was filtered through 105 µm pore nylon mesh (Component Supply Co, U-CMN-105-A) prior to carrying out flow cytometric analysis using an LSRFortessa instrument (BD Biosciences).The following antibodies and dilutions were used to stain the fibroblast cells: CD45 (1:100), CD31 (1:100), OSMR (1:100), PDPN (1:100). $5 \times 10^3$ counting beads (Spherotech, ACRFP-100-3) were added to each sample to enable quantification of cell numbers, and results were analyzed using FlowJo V10 software (Flowjo, LLC). A representative gating strategy is provided in **Supplementary Data Figure 1**.

**Analysis of intestinal barrier integrity**

To assess paracellular epithelial leakiness across the intestinal barrier in Healthy and IBD Colon Chips, Cascade Blue (0.59 or 3 kilodaltons) (C3239, D7132; Invitrogen, ) was passed through the apical channel at 50 μg/mL. Initial and final outflows of both channels were collected, and luminescence was measured according to the manufacturer's protocol. Permeability was calculated according to the following equation:

$$P_{app} = \frac{\left(\frac{dQ}{dt}\right)}{AdC} \qquad (1)$$

where A is the total area of diffusion, dC is the average concentration gradient and dQ/dt is molecular flux, as previously described[1].

**Transcriptomic Analysis**

RNA was extracted from Colon Chips at two weeks of culture, after formation of a functional epithelial-stromal interface with an overlying mucus layer. Cells in apical and basal channels were harvested separately with RLT lysis buffer from RNeasy Mini Kit (74106; Qiagen, Hilden, Germany), as previously described[3]. Channels were washed with 100 µL DPBS three times and then fibroblasts were lysed by quickly pressing and releasing the micropipette plunger into the input port of the lower channel at least four times. Lysates were collected in 1.5 mL tubes and stored immediately at -80°C for RNA sequencing analysis. Subsequently, epithelial cell lysates were collected and processed in a similar manner. High-grade RNA (RIN > 8) was prepared for transcriptomic analysis (Agilent Nano Kit 5067-1511 (Agilent 2100 Bioanalyzer, Agilent Technologies) and mRNA sequencing was performed by Azenta Life Sciences (Burlington, MA) via polyA selection using an Illumina HiSeq for 150 bp paired-end reads. Trimmomatic v.0.36 was used to remove possible adapter sequences and nucleotides with poor quality from sequence reads. STAR aligner v.2.5.2b was then used to map the trimmed reads to the *Homo sapiens* GRCh38 with ERCC genes reference genome available on ENSEMBL. Unique gene hit counts were calculated using feature counts from the Subread package v.1.5.2. Only unique reads that fell within exon regions were counted. For plotting of sample expression levels, gene counts were CPM normalized using edgeR v4.2.1[4].

Before differential expression analysis, genes were filtered to include only genes with at least 3 reads counted in at least 20% of samples in any group. Differential expression analysis was then performed with the DESeq2 R package[5], which tests for differential expression based on a model using the negative binomial distribution, and a sequencing batch was included in the design as a covariate. The false discovery rate (FDR) method was applied for multiple testing correction[6]. Gene set enrichment analysis (GSEA) was using the fgsea R package and the fgseaMultilevel() function[7]. The $\log_2$ fold change from the differential expression comparison was used to rank genes. C5: Gene Ontology gene sets - biological process gene set, C2: Canonical pathways – REACTOME, and C2: Canonical pathways - KEGG collections from the Molecular Signatures Database (MSigDB)[8,9] was curated using the msigdbr R package. Prior to running GSEA, the list of gene sets was filtered to include only gene sets with between 5 and 1000 genes. Differential expression and GSEA were performed using Pluto (https://pluto.bio).

The webapp g:Profiler[10] was used to perform functional enrichment analysis of differentially expressed genes. For each comparison, upregulated genes (p-adjusted < 0.05; log2fold change > 1) and downregulated genes (p-adjusted < 0.05; log2fold change < 1) were separately queried using g:Profiler across all data sources. The statistical data scope included only annotated genes and the g:SCS method was used for computing multiple testing corrections for p-values at a threshold of $p<0.05$. The R package ggplot2 (v3.5.1)[11] was used to generate dot plots of gene ontology terms from g:Profiler.

**Atomic Force Microscopy**

Atomic force microscopy (AFM) experiments were performed on different positions of unfixed, snap-frozen cryosections (20 μm thick) of patient intestinal tissue samples adhered to glass and immersed in PBS. AFM nanoindentation tests were performed using a Leica LAS X widefield system equipped with Bruker JPK Nanowizard 4a with a 5 μm silicon nitride cantilever (SAA-SPH-5UM), and all sectioned samples were maintained in PBS during experiments. Cantilevers were thermally calibrated before sample testing, with nominal values of 148-215 pN/nm. The positions for AFM scanning on the unstained sections were chosen based on the image of an H&E stained reference section. 2D force maps were taken in 20 x 20 or 40 x 40 μm square grids with 16-32 samples points per axial dimension. AFM measurements were made using a cantilever deflection setpoint of 10 nN and an indentation rate of 20 μm/s to measure elastic properties and minimize viscoelastic effects. Force-indentation curves were analyzed using a modified Hertz model for contact mechanics of spherical elastic bodies, where Poisson's law was assumed to be 0.4, as described[12]. To obtain Young's modulus, force-indentation curves were fitted according to Equation 2:

$$F = \frac{4E}{3}\sqrt{R\delta^3} \qquad (2)$$

Where E is apparent Young's modulus, μ is Poisson's ratio R is tip radius, F is the force and v is the indentation distance into the material.[25-28]

**Immune Cell Recruitment Assay**

Chips were removed from the Pods that hold the Organ Chip devices (Emulate Inc.), and 25 μl of the PBMC suspension were added to the lower channel. The chips were flipped and placed back in the incubator at 37°C to allow adhesion for 2 hours. Afterward, the chips were flipped back and 100 μl of culture medium was perfused through the lower channel to remove unattached cells. The chips were then reinserted into the Pods, placed in the Zoe culture instrument (Emulate Inc.), and flow and cyclic strain were resumed. 24 hours post-inoculation, supernatants were collected, and the chips were immediately imaged using an ECHO fluorescence microscope. Cell counting was performed with the Cell Counter plugin in ImageJ.

**Whole Genome Sequencing**

In the first batch of shallow whole genome sequencing, DNA was fragmented by sonication using a Diagenode Bioruptor (Invitrogen; 30 s ON/90 s OFF, high potency, 10 cycles). Fragmented DNA was checked using Tapestation and D1000HS tapes and reagents (Agilent Technologies, Santa Clara, CA, USA). Between 0.5-5ng of sonicated DNA was used for preparation of the libraries for whole genome shallow pass sequencing using the NEBNext Ultra II FS DNA Library Prep Kit (New England BioLabs, Ipswich, MA) according to the manufacturer instructions and using 13 cycles of library amplification. Libraries were cleaned with Clean NGS magnetic beads (GC Biotech), checked with Agilent TapeStation and D1000 tapes and reagents (Agilent Technologies, Santa Clara, CA, USA), and then equimolarly pooled. Sequencing was performed on Illumina's NextSeq 500 platform (High Output Run, 75bp paired-end reads), generating a mean depth of 0.34x (range 0.14x to 0.96x). The second batch of whole genome shallow pass sequencing was performed by Azenta Life Sciences (Burlington, MA) using an Illumina HiSeq for 150 bp paired-end reads.

**Data and code availability**

Data that support the findings of this study are available within the paper and its Supplementary Information files. The RNA sequencing data have been deposited in the Gene Expression Omnibus (GEO) with the accession code GSE277964. Any other material including the R code used for data analysis is available upon reasonable request.

**Extended Data Figure 1. Establishment of Healthy and IBD biobanks and Colon Chips**

**A)** Photograph of healthy, Crohn's and UC colon tissues used to obtain cell resources. Unaffected regions from healthy colon and inflamed regions in IBD tissues were used to obtain mucosa containing epithelium and stroma **B)** Establishment of epithelial organoid and fibroblast biobanks from patient resections. Digested mucosal tissues were split between organoids in Matrigel and 2D culture flasks for fibroblast culture. After day 7, cells were either passaged or cryostored to create the biobank. (bar, 200 μm). **C)** Representative transmitted light images of whole healthy and IBD Colon chips 11 days after seeding**.** A functional epithelial monolayer with crypt-like structures were formed in the apical channel of all chips. The number of crypt-like structures were lower in UC chips. (bar, 1 mm). **D)** Immunofluoresence vertical cross-section micrographs of healthy and IBD Colon chips showing the crypt-like epithelial structures in the Colon Chips. Magenta, phalloidin; cyan: DAPI ; Dashed white line, upper surface of chip membrane; dashed green line, upper luminal boundary of the crypt (bar, 100 μm)**.**

**Extended Data Figure 2: Characterization of IBD fibroblasts. A)** Histogram of low and high OSMR expression in CD45- and CD31- IBD fibroblasts. **B)** Podoplanin (PDPN) expression in CD45- and CD31- fibroblasts from IBD patients with low and high OSMR expression. High OSMR expression is correlated with high PDPN expression in IBD patients that contain inflammatory fibroblast subsets. 2D cell culture data was established with cells from 3 healthy (green), 2 Crohn's (magenta), and 2 UC (cyan) patient donors, with each symbol represents a chip created with cells from a different patient. All data represent mean ± SD; *p* values from student's t-test are shown in the figure. **C)** On-chip characterization of markers associated with fibroblast activation (Bar 200 μm) **D)** Vimentin and α-SMA were found to be expressed higher in IBD phenotype. Each symbol represents and individual Healthy or IBD patient. Chips were created with cells from 3 healthy (green), 2 Crohn's (magenta), and 2 UC (cyan) patient donors, with each symbol represents a chip created with cells from a different patient. All data represent mean ± SD; *p* values from student's t-test are shown in the figure. Source data and statistical tests are provided as a Source Data file.

**Extended Data Figure 3: Regulation of genes in epithelium and stroma from healthy and IBD Colon Chips. A)** Volcano plot of differentially expressed genes in A) Crohn's and B) UC chip epithelium compared to Healthy chip. C) Crohn's and D) UC chip fibroblast compared to Healthy chip. Chips were created with cells from 3 healthy, 2 Crohn's, and 2 UC patient donors.

**Extended Data Figure 4: Regulation of pathways in epithelium and stroma from healthy and IBD Colon Chips. A)** Dot plot of pathways in A) UC and B) Crohn's chip epithelium compared to Healthy chip. C) UC and D) Crohn's chip fibroblast compared to Healthy chip. Chips were created with cells from 3 healthy, 2 Crohn's, and 2 UC patient donors.

**Extended Data Figure 5**: **Micromechanical properties of healthy and IBD patient mucosa**. **A)** Transmitted light showing the AFM head on sectioned colon tissue from a patient. (Bar 100 μm). **B)** Contours showing that stromal Young's modulus is higher in IBD patients compared to Healthy. The white line shows the basement membrane. **C)** Representative sample atomic force microscopy curves of indentation and retraction of the cantilever from healthy and IBD patient stroma section on the microscope slide. **D)** Violin plot of measurements showing that stiffness of colon stroma of IBD patients was higher than healthy. Numbers indicate *P* values between compared groups, as determined by two-tailed Student's *t*-test. (n=2 Healthy and n=1 Crohn's and n=1 UC patient tissues). Source data and statistical tests are provided as a Source Data file.

**Extended Data Figure 6: Peristalsis-like cyclic strain can alter mucus and ECM production and their associated biological processes and metabolism. A)** Representative side-view images of whole Healthy or IBD Chips with (+) and without (-) exposure to mechanical deformations visualizing mucus layer accumulation (white diffuse material) in Healthy Chips using dark-field microscopy (bar, 1 mm). **B)** Peristalsis-like cyclic strain on IBD Colon Chips activates pathways and biological processes associated with fibrillar collagen production and organization in IBD fibroblasts (Chips were created with cells from, 2 Crohn's, and 2 UC patient donors). **C)** IBD Colon Chips exhibit deficient metabolization associated gene expression. Multiple cytochromeP450 subunits and families were found to be expressed lower in epithelium of IBD Colon Chip. Epithelium of healthy Colon Chips exposed to peristalsis-like cyclic strain can increase expression of majority of CYPs but does not improve IBD Colon Chips. Chips were created with cells from 3 healthy, 2 Crohn's, and 2 UC patient donors. **D)** Heatmaps showing relative expression of mucus production-associated genes when chips were stimulated with peristalsis-like cyclic strain versus non-stimulated. Cyclic strain increases expression of multiple mucins in epithelium of healthy chips but only limited amounts in IBD chips. (n = 3 healthy, 1 Crohn's and 2 UC). Source data and statistical tests are provided as a Source Data file.

**Extended Data Figure 7: Peristalsis-like cyclic strain drives inflammation in healthy and IBD Colon Chips. A)** Cyclic strain increased permeability of healthy and IBD epithelium to 3 kDa cascade blue particles. Chips were created with cells from 3 healthy (green), 2 Crohn's (magenta), and 2 UC (cyan) patient donors, with each symbol represents a chip created with cells from a different patient. All data represent mean $\pm$ SD; *p* values from two-way ANOVA test are shown in the figure. **B)** GSEA plot showing peristalsis-like cyclic strain activated the inflammatory response of epithelium on Healthy and IBD Colon Chips. (n=2 Crohn's and n=2 UC chips.). Source data and statistical tests are provided as a Source Data file.

**Extended Data Figure 8: Regulation of genes in epithelium and stroma from healthy and IBD Colon Chips under the peristalsis-like cyclic strain. A)** Volcano plot of differentially expressed genes in IBD epithelium and **B)** fibroblast that were exposed to peristalsis-like cyclic strain compared to chips cultured without strain. **C)** Volcano plot of differentially expressed genes in healthy epithelium and **D)** fibroblast that were exposed to peristalsis-like cyclic strain compared to chips cultured without strain. Chips were created with cells from 3 healthy, 2 Crohn's, and 2 UC patient donors.

**Extended Data Figure 9: GSEA analysis showing peristalsis-like cyclic activates pathways associated with inflammation and disease progression in A) epithelium and B) fibroblast of IBD Colon Chips.** Chips were created with cells from, 2 Crohn's, and 2 UC patient donors.

**Extended Data Figure 10: Second harmonic microscopic images of different fibrillar collagen (green) structures produced by IBD fibroblasts after being exposed to E2 + MPA + PAH. (bar, 200 μm).**

**Extended Data Figure 11: IBD fibroblasts and cyclic strain can partially modulate inflammation and expression of adhesion molecules. A)** Inflammatory cytokine and chemokine production in Healthy epithelium under the influence of Healthy and IBD fibroblasts. The fibroblast phenotype can increase inflammation in Healthy Colon Chips. Chips were created with cells from 2 healthy (green), 2 Crohn's, and 2 UC patient donors, with each symbol represents a chip created with cells from a different patient. All data represent mean $\pm$ SD; $p$ values from one-way ANOVA test are shown in the figure. **B)** Inflammatory cytokines produced by IBD epithelium that were not dependent on fibroblast phenotype in Colon Chip. Chips were created with cells from 2 healthy, 1 Crohn's (magenta), and 2 UC (cyan) patient donors, with each symbol represents a chip created with cells from a different patient. All data represent mean $\pm$ SD; $p$ values from one-way ANOVA test are shown in the figure. **C**) Higher expression of adhesion molecules in epithelium and fibroblasts was found in healthy and IBD Colon Chips. Peristalsis-like cyclic strain can increase expression of adhesion molecules only in IBD colon chips' epithelium. Chips were created with cells from 3 healthy, 2 Crohn's, and 2 UC patient donors. **D)** IL-6 and IL-8 production in healthy and IBD colon chips with PBMCs. IBD Colon Chips still retain high IL-6 and IL-8 production compared to healthy colon chips with PBMCs.

Chips were created with cells from 2 healthy (green), 1 Crohn's (magenta), and 1 UC (cyan) patient donors, with each symbol represents a chip created with cells from a different patient. All data represent mean $\pm$ SD; *p* values from one-way ANOVA test are shown in the figure.

**Extended Data Figure 12: Expression of carcinogenesis markers in healthy and IBD epithelium when co-cultured with different sources of fibroblasts and after being exposed to ENU. A) Confocal micrographs of patient tissues stained for DAPI (gray) and** early stage CRC marker CEACAM5 (green), which is expressed only in CRC tissues but not in healthy and IBD tissues. (bar 50 μm). **B)** CEACAM5 staining of healthy and IBD chips when cultured without fibroblast or with fibroblast from healthy and IBD tissues showing it is not expressed in healthy and IBD epithelium without ENU exposure (bar 50 μm) **C**) Quantification of CEACAM5 in healthy and IBD epithelium after 3 weeks of exposure to ENU. **D)** Quantification of protein staining intensity and morphology reveal that ENU exposure decreases E-cadherin levels and increases nuclear localization of β-catenin in IBD patient-derived Colon Chips compared to Healthy Chips.

**Supplementary Data Figure 1: Representative flow cytometry gating scheme to quantify OSMR and PDPN expression of IBD fibroblasts.**
